# Nasal transcriptomics characterize T-helper (T)2-low asthma endotypes in a study of Puerto Rican and Dominican adults living in the U.S.

**DOI:** 10.64898/2026.09.14.26363037

**Authors:** Sheng Yang, Yueh-Ying Han, Soheil Farid Azar, Amber Pirzada, Franziska J. Rosser, Golda Hudes, Robert Kaplan, Wei Chen, Carmen R. Isasi, Rasika A. Mathias, Juan C. Celedón

**Author notes:** Corresponding author: Juan C. Celedón, MD, DrPH, Division of Pediatric Pulmonary Medicine, UMPC Children’s Hospital of Pittsburgh, 4401 Penn Avenue, Pittsburgh, PA 15224.

## Abstract

**IMPORTANCE:** CD4+ T helper (T) 1, T2 and T17 cells have helped identify asthma endotypes in predominantly non-Hispanic White adults. Identifying asthma endotypes in diverse subgroups should help develop personalized medicine in asthma.

**OBJECTIVE:** To characterize asthma endotypes and their association with selected variables and asthma severity/control in adults.

**DESIGN, SETTING, AND PARTICIPANTS:** Cross-sectional and longitudinal analyses of nasal samples from the SOL-Asthma study of Puerto Rican and Dominican adults with (cases, n=313) and without (controls, n=371) asthma in the Bronx (NY) and Chicago (IL), with replication of selected findings in 376 adults of African ancestry in CAAPA.

**MAIN OUTCOMES AND MEASURES:** Our primary outcome was asthma endotypes identified through nasal expression of “signature genes” for 3 T2, 5 T1, and 5 T17 pathways. Multinomial logistic regression was used for the analysis of asthma endotypes (with controls as the reference outcome category) at a baseline visit, as well as for the analysis of severe asthma exacerbations (SAEs) at baseline and during a 1-year follow-up. We attempted to replicate the identified profiles in CAAPA.

**RESULTS:** In SOL-Asthma, cases were more likely to be female, Puerto Rican, and current/former smokers, and to have physician-diagnosed COPD than controls. Five transcriptomic profiles were identified in cases: T2^HIGH^ (n=16 [5.1%]), T1^HIGH^ (n=71 [22.7%]), T17^HIGH^ (n=96 [30.7%]), T1^HIGH^/T17^HIGH^ (n=53 [16.9%]), and T2^LOW^/T1^LOW^/T17^LOW^(n=77 [24.6%]). In a multivariable analysis, Puerto Ricans were more likely to have a T17^HIGH^ profile than Dominicans (odds ratio [OR]=2.48, 95% confidence interval [CI]=1.08-5.74) and female sex was associated with T1^HIGH^ (OR=3.44, 95% CI=1.33-8.88), T1^HIGH^/T17^HIGH^and T2^LOW^/T1^LOW^/T17^LOW^ profiles, while COPD was associated with profiles other than T2_HIGH_ or T1_HIGH_ The Ti_HIGH_ profile was associated with ≥1 SAE in the year prior to baseline and during the one-year follow-up (OR=4.31, 95% CI=1.26-14.77). Each profile had unique differentially expressed genes (DEGs), with substantial replication of the profile distribution and DEGs in CAAPA.

**CONCLUSIONS AND RELEVANCE:** Nasal transcriptomics identified profiles corresponding to five asthma endotypes in a cohort of Puerto Rican and Dominican adults. Puerto Rican background, female sex, and physician-diagnosed COPD were associated with non-T2^HIGH^ (T2^LOW^) asthma endotypes, and T1^HIGH^ asthma was strongly associated with SAEs.

**KEY POINTS:** *Question:* Can nasal transcriptomics identify asthma endotypes and associated variables and outcomes in high-risk adults?

*Findings:* Five transcriptomic profiles were identified in Puerto Rican and Dominican adults: T helper (T)2^HIGH^, T1^HIGH^, T17^HIGH^, T1^HIGH^/T17^HIGH^, and T2^LOW^/T1^LOW^/T17^LOW^. Puerto Ricans were more likely to have a T17^HIGH^ profile than Dominicans and female sex was associated with T1^HIGH^, T1^HIGH^/T17^HIGH^ and T2^LOW^/T1^LOW^/T17^LOW^ profiles, while COPD was associated with profiles other than T2^HIGH^ or T1^HIGH^. The T1^HIGH^ profile was strongly associated with severe asthma exacerbations.

*Meaning:* Women and Puerto Ricans were more likely to have T2^LOW^ asthma endotypes, for which there is no specific treatment.

## INTRODUCTION

Asthma is a leading cause of morbidity worldwide.^1,2^ In the United States (U.S.), Puerto Ricans and non-Hispanic Blacks are heavily affected with asthma. Among adults in the Hispanic Community Health Study/Study of Latinos (HCHS/SOL), the prevalence of current asthma was higher in Puerto Ricans (21.9%) than in Dominicans (6.7%) and Mexicans (3.1%).^3^ In another U.S. study, the age-specific standardized mortality rates for asthma (per 100,000 people) were much higher in non-Hispanic Blacks (28.76) and Puerto Ricans (25.42) than in non-Hispanic Whites (8.61).^4^

Asthma is a heterogenous condition. Studies of gene expression in bronchial epithelium and sputum from predominantly non-Hispanic White adults with asthma have identified endotypes with variable treatment response. T helper (T)2-high asthma is characterized by eosinophilic airway inflammation and high interleukin (IL)-4 and IL-13 serum levels; T17-high asthma is characterized by neutrophilic airway inflammation, high IL-17 serum levels, and corticosteroid resistance; and T1-high asthma has been linked to viral-driven asthma exacerbations through IFN-γ and STAT1 signaling.^5–7^ While severe T2-high asthma can be treated with monoclonal antibodies, there is no specific treatment for non-T2 high (T2-low) asthma endotypes.^8^

We previously showed that nasal expression of “signature genes” for T2, T1 and T17 immune responses identifies transcriptomic profiles in youths corresponding to the asthma endotypes identified in adults.^9^ Moreover, T2-low asthma endotypes were more common than T2-high asthma in cross-sectional studies of predominantly Puerto Rican and non-Hispanic Black children and adolescents.

We hypothesized that T2-low asthma endotypes would be common and associated with worse asthma outcomes in Puerto Rican and Dominican adults. To test this hypothesis, we identified nasal transcriptomic profiles corresponding to asthma endotypes and examined whether T2-low asthma endotypes are associated with worse disease outcomes in a cohort of Puerto Rican and Dominican adults in HCHS/SOL. We then attempted to replicate some of our findings in adults of African ancestry who participated in the Consortium on Asthma among African ancestry Populations in the Americas (CAAPA).

## METHODS

Please also see the **Online Supplement (OS)**.

### Study population

HCHS/SOL is a population-based cohort study of 16,415 adults aged 18 to 74 years who self-identified as Hispanic/Latino at screening visits of households in Chicago (IL), Miami (FL), Bronx (NY) and San Diego (CA).^10^ A baseline visit was conducted between March 2008 and June 2011^10^, with yearly telephone assessments afterwards. The sample design and cohort selection were previously described.^11^ In brief, a stratified two-stage probability sample of household addresses was selected in each center to provide a representative sample of the target Hispanic population. The study oversampled persons aged 45-74 years (n=9,714, 59.2%) to increase the proportion of target outcomes, and the baseline visit included questionnaires, spirometry, and collection of blood samples.^10,12^ Spirometry was performed using the SensorMedics model 1022 dry-rolling seal volume spirometer (SensorMedics/Viasys, Yorba Linda, CA), following American Thoracic Society (ATS) recommendations^4^. The study was approved by the Institutional Review Boards (IRBs) at the Data Coordinating Center (University of North Carolina) and at each center, and all participants gave written informed consent.

An ancillary study (SOL-Asthma) was conducted in 748 Puerto Rican and Dominican adults with (cases, *n*=374) and without (controls, *n*=374) asthma. Subjects who consented to be contacted for ancillary studies were recruited at the Bronx and Chicago centers, which enrolled most Puerto Rican and Dominican participants in HCHS/SOL, between April 2022 and May 2025. Eligibility criteria included self-reported Puerto Rican or Dominican background and ability to provide written informed consent. Asthma was defined by a positive response to all following questions: “Have you ever had asthma?”, “Was it diagnosed by a doctor or other health care professional?”, and “Do you still have it?” Controls had never had asthma. A baseline visit included questionnaires on respiratory health and the Asthma Control Test (ACT),^13^ spirometry, and collection of nasal samples (as previously described).^14^ Spirometry was conducted with an Easy One spirometer (ndd Med. Technol., Andover, MA) following ATS/ERS recommendations,^15,16^ and testing was repeated 15’ after two puffs of an albuterol MDI. A year after the baseline visit, a follow-up phone call including a brief questionnaire on respiratory health and the ACT was completed in cases.

Of the 748 participants in SOL-Asthma, 684 were included in this analysis: 313 cases with nasal RNA-seq data and 371 controls (**Figure 1, Panel A**). SOL-Asthma was approved by the IRBs of the Albert Einstein College of Medicine and the University of Illinois at Chicago.

**Figure 1.**
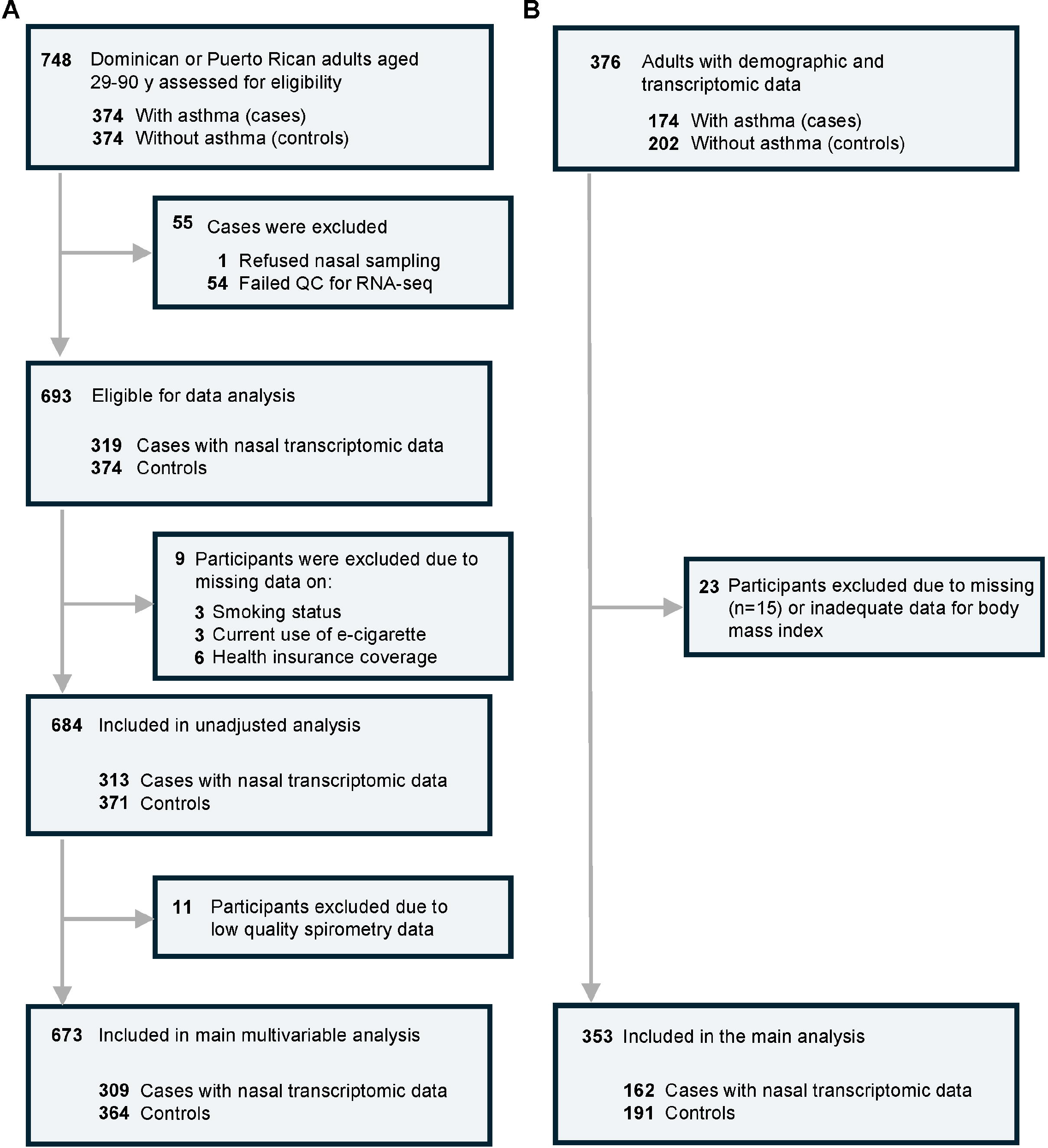
Flowchart for selection of participants in the Study of Latinos (SOL)-Asthma Study (Panel A) and the Consortium on Asthma among African ancestry Populations in the Americas (CAAPA) for the current study (Panel B).

RNA-seq was conducted at the Genomics Core of the University of Pittsburgh using RNA from nasal samples. To remove cytoplasmic and mitochondrial ribosomal RNA, library preparation was done using TruSeq Stranded Total RNA Library Prep Kit with TruSeq Stranded mRNA (Illumina, San Diego, CA). Libraries were run on the Illumina NextSeq 500 platform with paired-end 75-100 cycles and with 80-100 million reads per sample; reads were aligned to reference human genome (GRCh38) and Transcripts Per Million (TPM) were used as proxy for gene expression level.^18^

As in prior work, nasal expression of “signature genes” for T2 (periostin [*POSTN*], serpin family B member 2 [*SERPINB2*], and chloride channel, calcium-activated, family member 1 [*CLCA1*])^19^, T1 (signal transducer and activator of transcription 1 [*STAT1*], interferon-gamma [*IFNG*], CD8 subunit alpha [*CD8A*], CD8 subunit beta [*CD8B*], and C-X-C motif chemokine ligand 10 [*CXCL10*])^5^, and T17 (C-X-C motif chemokine ligand 1 [*CXCL1*], 2 [*CXCL2*] and 3 [*CXCL3*]; colony-stimulating factor 3 [*CSF3*], and interleukin-8 [*IL8]*)^20^ immune pathways was first normalized to TPM values and then transformed to a log_2_ scale after adding a constant value (0.1).

K-means clustering analysis was performed in cases based on expression of the “signature genes”, testing a number of clusters (k) ranging from three to eight. We used NbClust (version 3.0.1) in R to evaluate 30 clustering indices (e.g., silhouette width) by majority rule.^21^ Eight of 23 applicable indices supported k=5 as the optimal choice, resulting in five transcriptomic profiles (**eFigure 1**), as follows: upregulated expression of T2 genes only (“T2-high”), upregulated expression of T1 genes only (“T1-high), upregulated expression of T17 genes only (“T17-high), upregulated expression of T1 and T17 genes (“T1-high/T17-high”), and no upregulated expression of T2, T1, or T17 genes (“T2-low/T1-low/T17-low”). The K-means clustering analysis was validated using Gaussian mixture modeling (GMM), based on expression of the same signature genes. Agreement between K-means and GMM classifications was quantified using the Adjusted Rand Index (ARI).

### Statistical analysis

Our primary outcomes in SOL-Asthma were the nasal transcriptomic profiles corresponding to asthma endotypes. We compared key characteristics between controls and each asthma endotype using ANOVA or the Kruskal-Wallis test for continuous variables and Chi-squared or Fisher’s exact tests for categorical variables. Given the complex sampling design of HCHS/SOL, all multivariable analyses incorporated sampling weights, stratification, and clustering to obtain unbiased estimates and standard errors. Multinomial logistic regression was used for the analysis of asthma endotypes, with controls as the reference outcome category.^22^ All models included age, sex, body mass index (BMI), Hispanic/Latino background (Puerto Rican vs. Dominican), study site (Bronx vs. Chicago), birthplace in the 50 U.S. states or Washington, D.C., having employer-based or private health insurance, smoking status (never, former, or current), pack-years of smoking, physician-diagnosed chronic obstructive pulmonary disease (hereafter, COPD), percent (%) predicted pre-bronchodilator (BD) forced expiratory volume in one second (FEV1), and pre-BD FEV1/forced vital capacity (FVC) ratio. Predicted values for FEV1 and FVC were calculated using Global Lung Function Initiative 2022 equations accounting for age, sex, and height.^23^

To reduce misclassification of COPD as asthma, we conducted a sensitivity analysis excluding current smokers and former smokers with ≥10 pack-years of smoking and participants with COPD. Further, we performed a sensitivity analysis adjusting for indicators of asthma severity/control other than lung function: current use of inhaled corticosteroids (ICS) and oral corticosteroids (OCS), ACT score, and ≥1 severe asthma exacerbation (SAE, defined as ≥1 emergency department (ED) visit or ≥1 hospitalization for asthma) in the prior year. Moreover, we conducted a sensitivity analysis adjusting for principal components (PCs) instead of Hispanic/Latino background in 580 participants with genome-wide (GW) genotypic data.

Using multivariable logistic regression with the T2^LOW^/T1^LOW^/T17^LOW^ endotype as the reference outcome category, we conducted prospective analyses of asthma endotypes at baseline and: ≥1 SAE and an ACT score decline ≥3 points during the 1-year follow-up in SOL-Asthma. This analysis was adjusted for ICS use and (for ACT score decline) baseline ACT score, in addition to the covariates included in the cross-sectional analysis. We also conducted a retrospective multivariable linear regression analysis of asthma endotypes and change in lung function measures between the baseline visit in HCHS-SOL and that in SOL-Asthma. This analysis was adjusted for BMI, Hispanic/Latino background, study site, birthplace in the 50 U.S. states or Washington, D.C., smoking status, pack-years of smoking, COPD, the corresponding lung function measure (e.g., FEV1) at the HCHS/SOL visit, and the time between visits.

Next, we conducted an analysis of differentially expressed genes (DEGs) for each asthma endotype (with T2^LOW^/T1^LOW^/T17^LOW^ asthma as reference), using negative binomial regression in DESeq2 (version 1.44.0)^24^ and adjusting for age, sex, BMI, Hispanic/Latino background, study site, and current ICS and OCS use. Genes were considered DEGs if the log2-fold change was >1 and the false discovery rate-adjusted P value (FDR-P) was <0.05. Enriched canonical pathways for upregulated DEGs for each asthma endotype (at FDR-P <0.01) were identified by ingenuity pathway analysis.^25^

All analyses were conducted in R (version 4.4.0).

### Replication analysis

We attempted to replicate some findings in SOL-Asthma in 376 CAAPA^17^ participants aged >18 years with (cases, *n*=174) and without (controls, *n*=202) current asthma. (**Figure 1, Panel B).** Using the same K-means clustering approach with 13 signature genes as in SOL-Asthma, cases were classified into five endotypes (**Figure 2, Panel B**). Since there was only one current smoker and persons with COPD were excluded from CAAPA, the multinomial logistic regression analysis of asthma endotypes (with controls as the reference outcome category) was adjusted for age, sex, BMI, former smoking, the first two PCs derived from GW genotypic data, study site, and %predicted pre-BD FEV1 and FEV1/FVC. As in SOL-Asthma, we analyzed DEGs between each endotype and T2^LOW^/T1^LOW^/T17^LOW^ asthma and conducted an enrichment analysis for upregulated genes.

**Figure 2.**
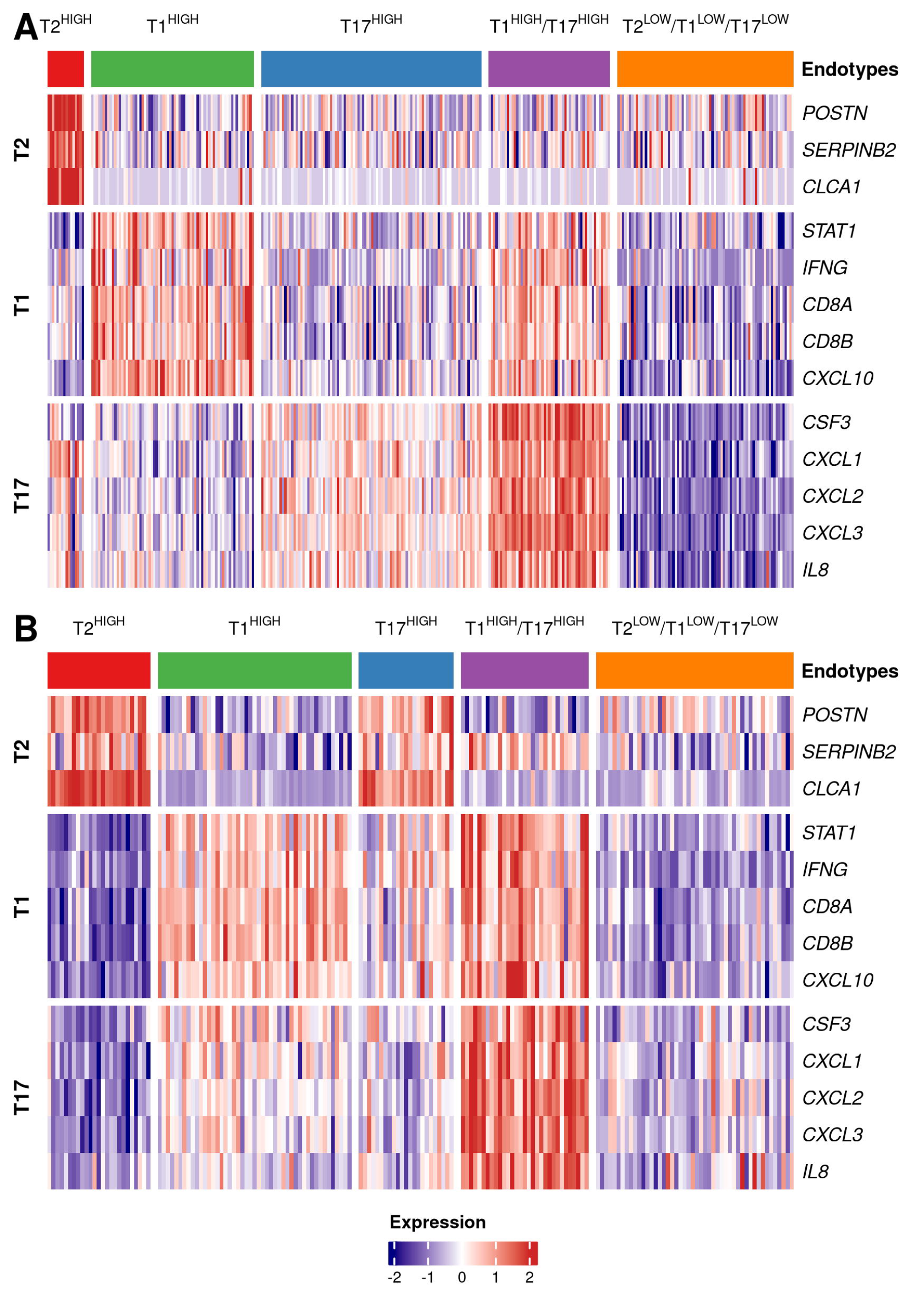
Heat map of five nasal transcriptomic profiles in the Study of Latinos (SOL)-Asthma Study (Panel A) and the Consortium on Asthma among African Ancestry Populations in the Americas (CAAPA, Panel B).

CAAPA was approved by the IRBs of the University of Colorado, Johns Hopkins University, University of Chicago, University of West Indies, University of Bahia, University of Ibadan, and the U.S. NIH.

## RESULTS

Please also see the **OS**.

Five transcriptomic profiles were identified in the K-means clustering analysis among cases in SOL-Asthma: T2^HIGH^(n=16 [5.1%]), T1^HIGH^ (n=71 [22.7%), T17^HIGH^ (n=96, [30.7%]), T1^HIGH^/T17^HIGH^ (n=53 [16.9%]), and T2^LOW^/T1^LOW^/T17^LOW^(n=77 [24.6%]) (**Figure 2, Panel A**). A sensitivity analysis using GMM yielded similar results (**eFigure 2**).

**Table 1** shows the main characteristics of the 684 SOL-Asthma participants, by asthma status and endotype. Compared with controls, cases with: 1) T2^HIGH^ asthma had higher total IgE and were more likely to be at the Bronx site and sensitized to ≥1 allergen; 2) T1_HIGH_ and T17^HIGH^ asthma were more likely to be Puerto Rican; 3) T17^HIGH^ asthma had higher BMI; 4) T1^HIGH^, T17^HIGH^ and T2^LOW^/T1^LOW^/T17^LOW^ asthma were more likely to be former smokers and had lower household income and higher pack-years of smoking; 5) T1^HIGH^ and T2^LOW^/T1^LOW^/T17^LOW^ asthma were more likely to be female; and 6) all endotypes other than T2^HIGH^ asthma were more likely to have COPD and parental asthma. Among cases, those with T1_HIGH_ asthma were most likely to have had ≥1 SAE in the prior year.

**Table 1.** Main characteristics of 684 participants in the Study of Latinos (SOL)-Asthma, by asthma status and asthma endotype.

| Characteristics | Without asthma<br>(controls,<br>*n*=371) | Asthma<br>(cases, *n*=313) |  |  |  |  |
| --- | --- | --- | --- | --- | --- | --- |
|  |  | T2<sup>HIGH</sup><br>(*n*=16) | T1<sup>HIGH</sup><br>(*n*=71) | T17<sup>HIGH</sup><br>(*n*=96) | T1<sup>HIGH</sup>/T17<sup>HIGH</sup><br>(*n*=53) | T2<sup>LOW</sup>/T1<sup>LOW</sup>/T17<sup>LOW</sup><br>(*n*=77) |
| Age (years) | 61.4 ± 12.4 | 56.7 ± 14.0 | 63.0 ± 12.3 | 62.9 ± 9.7 | 60.4 ± 12.8 | 62.0 ± 12.2 |
| Female sex | 243(65.5) | 13(81.2) | **60(84.5)\*** | 73(76.0) | 42(79.2) | **62(80.5)\*** |
| Body mass index (kg/m<sup>2</sup>) | 30.0 ± 5.8 | 30.1 ± 9.0 | 31.2 ± 6.5 | **32.4 ± 6.2\*** | 32.0 ± 6.8 | 31.1 ± 6.6 |
| Hispanic/Latino background |  |  |  |  |  |  |
| Dominican | 145(39.1) | 8(50.0) | 17(23.9) | 20(20.8) | 16(30.2) | 22(28.6) |
| Puerto Rican | 226(60.9) | 8(50.0) | **54(76.1)\*** | **76(79.2)\*** | 37(69.8) | 55(71.4) |
| Study site |  |  |  |  |  |  |
| Chicago, IL | 101(27.2) | 0(0.0) | 25(35.2) | 22(22.9) | 16(30.2) | 13(16.9) |
| Bronx, NY | 270(72.8) | **16(100.0)\*** | 46(64.8) | 74(77.1) | 37(69.8) | 64(83.1) |
| Educational attainment |  |  |  |  |  |  |
| No high school diploma or GED | 124(33.4) | 6(37.5) | 29(40.8) | 41(42.7) | 15(28.3) | 23(29.9) |
| At most a high school diploma or GED | 103(27.8) | 4(25.0) | 14(19.7) | 18(18.8) | 11(20.8) | 20(26.0) |
| Greater than high school (or GED) education | 144(38.8) | 6(37.5) | 28(39.4) | 37(38.5) | 27(50.9) | 34(44.2) |
| Born in the 50 U.S. states or D.C. | 96(25.9) | 7(43.8) | 23(32.4) | 26(27.1) | 17(32.1) | 26(33.8) |
| Age of immigration (years)¶ | 23.6 ± 13.5 | 24.7 ± 15.0 | 22.8 ± 11.8 | 20.2 ± 12.5 | 22.9 ± 14.3 | 22.6 ± 13.7 |
| Household income < 30,000/year | 211(59.9) | 8(53.3) | **56(81.2)\*** | **76(80.9)\*** | 35(70.0) | **54(73.0)\*** |
| Has employer-based or private health insurance coverage | 99(26.7) | 6(37.5) | 15(21.1) | 22(22.9) | 14(26.4) | 19(24.7) |
| Season of nasal sampling |  |  |  |  |  |  |
| September ~ November | 88(23.7) | 2(12.5) | 15(21.1) | 24(25.0) | 15(28.3) | 20(26.0) |
| December ~ February | 95(25.6) | 5(31.2) | 24(33.8) | 19(19.8) | 8(15.1) | 11(14.3) |
| March ~ May | 130(35.0) | 3(18.8) | 22(31.0) | 37(38.5) | 20(37.7) | 27(35.1) |
| June ~ August | 58(15.6) | 6(37.5) | 10(14.1) | 16(16.7) | 10(18.9) | 19(24.7) |
| Pack-years of smoking | 7.1 ± 21.2 | 3.1 ± 7.0 | **8.1 ± 15.2\*** | **10.1 ± 21.2\*** | 4.9 ± 12.1 | **10.8 ± 19.7\*** |
| Smoking status |  |  |  |  |  |  |
| Never | 235(63.3) | 10(62.5) | 31(43.7) | 50(52.1) | 36(67.9) | 30(39.0) |
| Former | 92(24.8) | 3(18.8) | **30(42.3)\*** | **37(38.5)\*** | 13(24.5) | **23(29.9)\*** |
| Current | 44(11.9) | 3(18.8) | **10(14.1)\*** | **9(9.4)\*** | 4(7.5) | **24(31.2)\*** |
| Current use of e-cigarettes | 4(1.1) | 1(6.2) | 2(2.8) | 1(1.0) | 1(1.9) | 1(1.3) |
| Physician-diagnosed COPD | 10(2.7) | 1(6.2) | **8(11.3)\*** | **12(12.5)\*** | **7(13.2)\*** | **8(10.4)\*** |
| Parental history of asthma | 68(20.5) | 5(33.3) | **29(45.3)\*** | **31(37.3)\*** | **22(50.0)\*** | **27(43.5)\*** |
| Asthma Control Test (ACT) score | -- | 21.0(17.5,24.5) | 20.0(17.0,24.0) | 22.0(17.5,25.0) | 23.0(18.0,25.0) | 21.0(18.0,24.0) |
| Current use of inhaled corticosteroids | -- | 4(25.0) | 24(34.3) | 35(36.5) | 13(24.5) | 21(27.3) |
| Current use of oral corticosteroids | -- | 6(37.5) | 14(19.7) | 21(21.9) | 12(22.6) | 18(23.4) |
| ≥1 emergency department visit or hospitalization for asthma, prior year | -- | 4(26.7) | **23(38.3)\*** | 14(17.5) | 6(13.6) | 9(14.1) |
| Blood eosinophil count (cells/μL)<sup>†</sup> | 200(100,200) | 200(200,400) | 200(100,200) | 200(100,200) | 100(100,300) | 100(100,200) |
| Total IgE (IU/ml)<sup>†</sup> | 48.0(19.0,134.0) | **218.5(93.0,413.0)\*** | 48.0(11.0,159.0) | 49.5(24.0,132.5) | 119.0(18.0,274) | 56.0(28.0,139.0) |
| ≥1 allergen-specific IgE<sup>†</sup> | 139(38.8) | **12(75.0)\*** | 34(48.6) | 44(46.3) | 27(50.9) | 32(42.1) |
| %predicted pre-bronchodilator FEV1 | 90.7 ± 16.1 | 92 ± 19.8 | 86.2 ± 17 | **82.3 ± 18.8\*** | **81.4 ± 21.8\*** | 85.9 ± 21.7 |
| %predicted pre-bronchodilator FVC | 93.2 ± 15.4 | 98.1 ± 20.9 | 92.2 ± 14.3 | **87.8 ± 15.9\*** | 87.6 ± 20.4 | 92.2 ± 17.8 |
| Pre-bronchodilator FEV1/FVC (%) | 77.5 ± 8.0 | 75.9 ± 8.2 | **74.5 ± 9.8\*** | **74.2 ± 10.0\*** | **74.3 ± 9.3\*** | **74 ± 11.7\*** |
| %predicted post-bronchodilator FEV1 | 93.2 ± 16.7 | 94.9 ± 15.5 | 90.2 ± 17.5 | **85.6 ± 17.6\*** | **87.3 ± 18.0\*** | 89.7 ± 21.9 |
| %predicted post-bronchodilator FVC | 92.6 ± 15.3 | 98.3 ± 17.1 | 93.2 ± 14.4 | 89.3 ± 14.8 | 90.9 ± 15.3 | 92.4 ± 15.8 |
| Post-bronchodilator FEV1/FVC (%) | 80.2 ± 7.9 | 78.0 ± 6.5 | **76.9 ± 10.0\*** | **76.1 ± 9.8\*** | **76.8 ± 9.5\*** | **76.9 ± 12.0\*** |

COPD= chronic obstructive pulmonary disease; GED= General Education Diploma; FEV1= forced expiratory volume in 1 second; FVC= forced vital capacity.
Data are shown as n (%) for categorical variables and as mean (standard deviation) for continuous variables, with the exception of: ACT score, blood eosinophils and neutrophils and total IgE (all shown as median [interquartile range]). Numbers may vary due to missingness.
¶In participants born outside the 50 U.S. states and D.C.
\*P<0.05 for comparison of participants in each asthma endotype versus controls or (for outcomes in cases only) T2<sup>LOW</sup>/T1<sup>LOW</sup>/T17<sup>LOW</sup> asthma.
<sup>†</sup>Measured in blood samples collected at the baseline visit of the Hispanic Community Health Study/ Study of Latinos (HCHS/SOL).

**Table 2** shows the results of the multivariable analysis of asthma endotypes. Female sex was significantly associated with 3.34-3.44 times increased odds of T1^HIGH^, T1^HIGH^/T17^HIGH^ and T2^LOW^/T1^LOW^/T17^LOW^ asthma and Puerto Rican background was significantly associated with 2.48 times increased odds of T17^HIGH^ asthma but 0.16 times decreased odds of T2^HIGH^ asthma. Moreover, former smoking was significantly associated with 3.29 times increased odds of T1^HIGH^ asthma and current smoking was significantly associated with 3.94 times increased odds of T2^LOW^/T1^LOW^/T17^LOW^ asthma; COPD was significantly associated with 3.44 to 9.7 times increased odds of T17^HIGH^, T1^HIGH^/T17^HIGH^, and T2^LOW^/T1^LOW^/T17^LOW^ asthma; and pre-BD FEV1/FVC was significantly and inversely associated with T1^HIGH^, T17^HIGH^, and T1^HIGH^/T17^HIGH^ asthma.

**Table 2.**
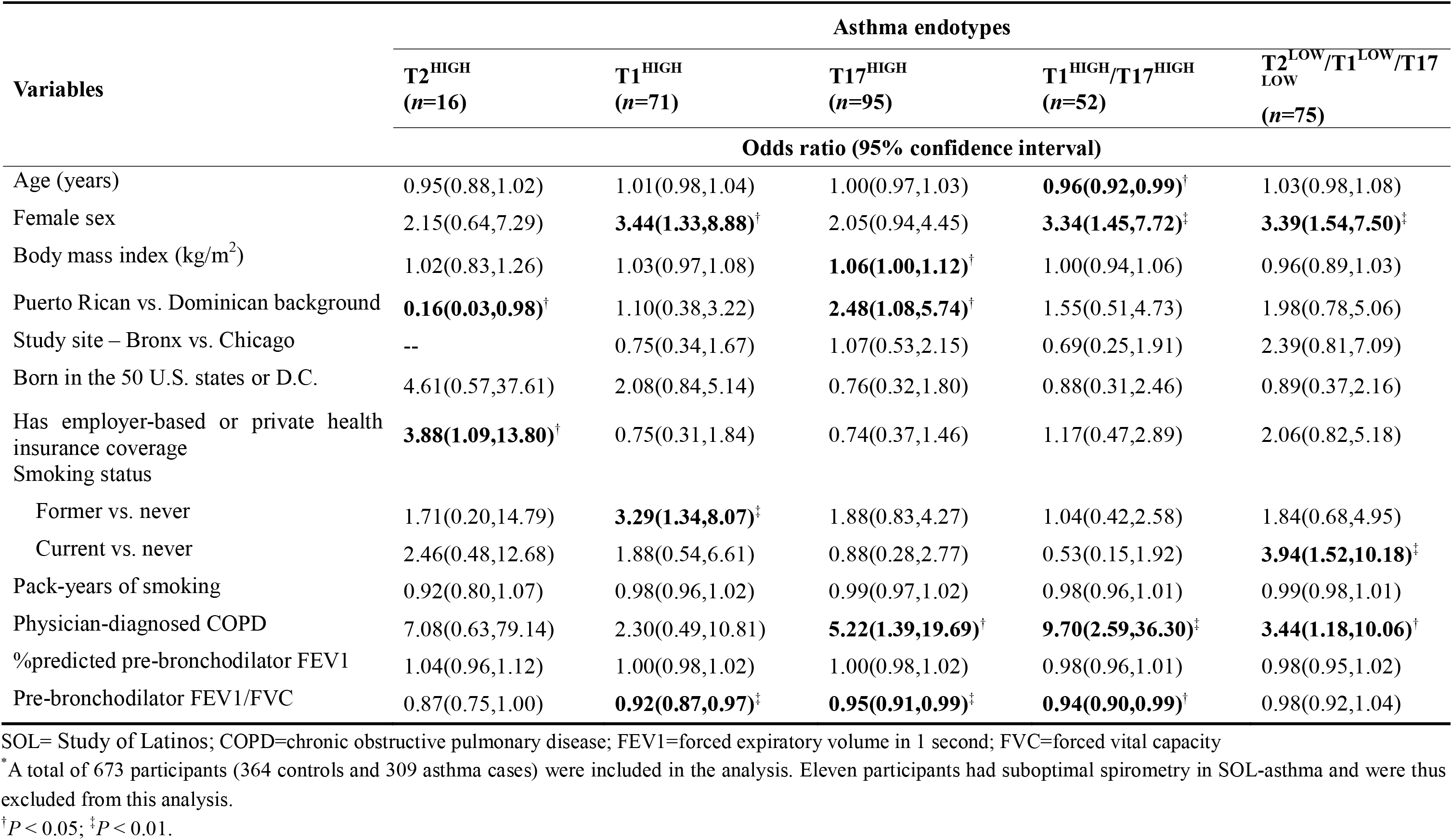
Multinomial logistic regression analysis of asthma endotypes in SOL-Asthma, with controls as the reference outcome category.

Next, we conducted a sensitivity analysis restricted to 472 participants without physician-diagnosed COPD who were never smokers or former smokers with <10 pack-years of smoking, obtaining similar findings despite smaller sample size (**eTable 1**). Further, we performed another sensitivity analysis adjusting for household income instead of health insurance coverage, obtaining similar results to those of the primary analysis while noting an association between low household income and T2^HIGH^ asthma (**eTable 2**). In another sensitivity analysis, we adjusted for PCs instead of Hispanic/Latino background in 580 participants with GW genotypic data, obtaining similar findings (**eTable 3**).

We then examined whether indicators of asthma control/severity differed by asthma endotype at baseline. In a multivariable analysis with T2^LOW^/T1^LOW^/T17^LOW^ asthma as the reference outcome category, cases with T1_HIGH_ asthma had 6.95 times significantly increased odds of ≥1 SAE in the previous year, adjusting for smoking, current use of OCS and ICS, and other covariates (**eTable 4**). We then conducted a prospective analysis of asthma endotypes and SAEs during the 1-year follow-up in SOL-Asthma (**Figure 3** and **eTable 5**). After adjustment for ICS use, smoking, and other covariates, cases with T1_HIGH_ asthma had fourfold significantly higher odds of ≥1 SAE during follow up than those with T2^LOW^/T1^LOW^/T17^LOW^ asthma. No asthma endotype was associated with ACT score decline (**eTable 6**).

**Figure 3.**
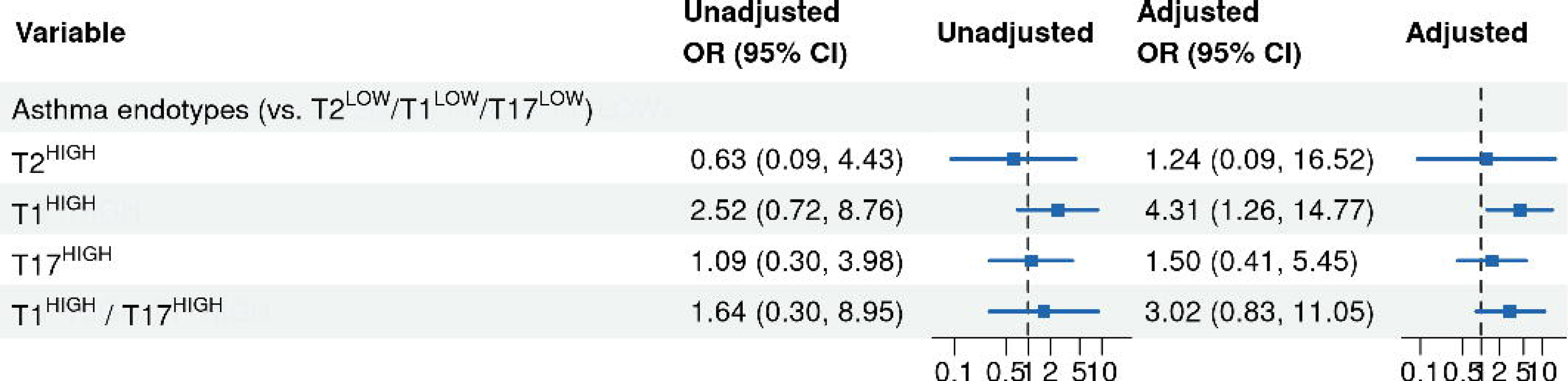
Forest plot for the results of the multinomial regression analysis of asthma endotypes and self-reported emergency department visits or hospitalizations for asthma during the 1-year follow up in SOL-Asthma. <u>Footnote</u>: OR=odds ratio and CI=confidence interval. The multivariable analysis was adjusted for age, sex, body mass index, Hispanic/Latino background (Puerto Rican vs. Dominican), study site (Bronx vs. Chicago), birthplace (born in 50 U.S. states/D.C.), physician-diagnosed COPD, percent predicted pre-bronchodilator FEVD, and pre-bronchodilator FEVD/FVC ratio. In this analysis, the reference group is T2^LOW^/T1^LOW^ /T17^LOW^ asthma. The dashed vertical line indicates OR = 1.

Next, we conducted a retrospective multivariable analysis of change in lung function measures between the HCHS/SOL baseline visit and that in SOL-Asthma in 597 participants (**eTable 7**). Compared with controls, participants with T1^HIGH^, T1^HIGH^/T17^HIGH^ and T2^LOW^/T1^LOW^/T17^LOW^ asthma had significant decrements of 1.99%-2.29% in FEV1/FVC. Further, COPD was significantly associated with a 4.15% reduction in FEV1/FVC.

**Figure 4** and **eFigure 3** show the results of the analyses of DEGs and pathway enrichment in SOL-Asthma. In these analyses, 892 genes were upregulated in T2^HIGH^ asthma, with strongest upregulation for *CLC, CCR3*, *CTSG*, *SIGLEC8*, and *HPGDS*. In T1^HIGH^ asthma, 387 genes (e.g., *CXCL9*, *GZMB*, and *STAT1*) were upregulated, while in T17^HIGH^ asthma 209 genes (e.g., *CXCL5*, *DEFB4A*, and *IL1B*) were upregulated. In T1^HIGH^/T17^HIGH^ asthma, 977 genes were upregulated, with highest upregulation for *CXCL5*, *IL1B*, *CSF3*, *FPR1*, and *FPR2*, consistent with concurrent activation of neutrophil recruitment and innate inflammatory signaling. **eFigure 4** shows a heatmap using an expanded signature gene panel derived from integrating WGCNA and DEG analyses, with similar findings to that using 13 signature genes.

**Figure 4.**
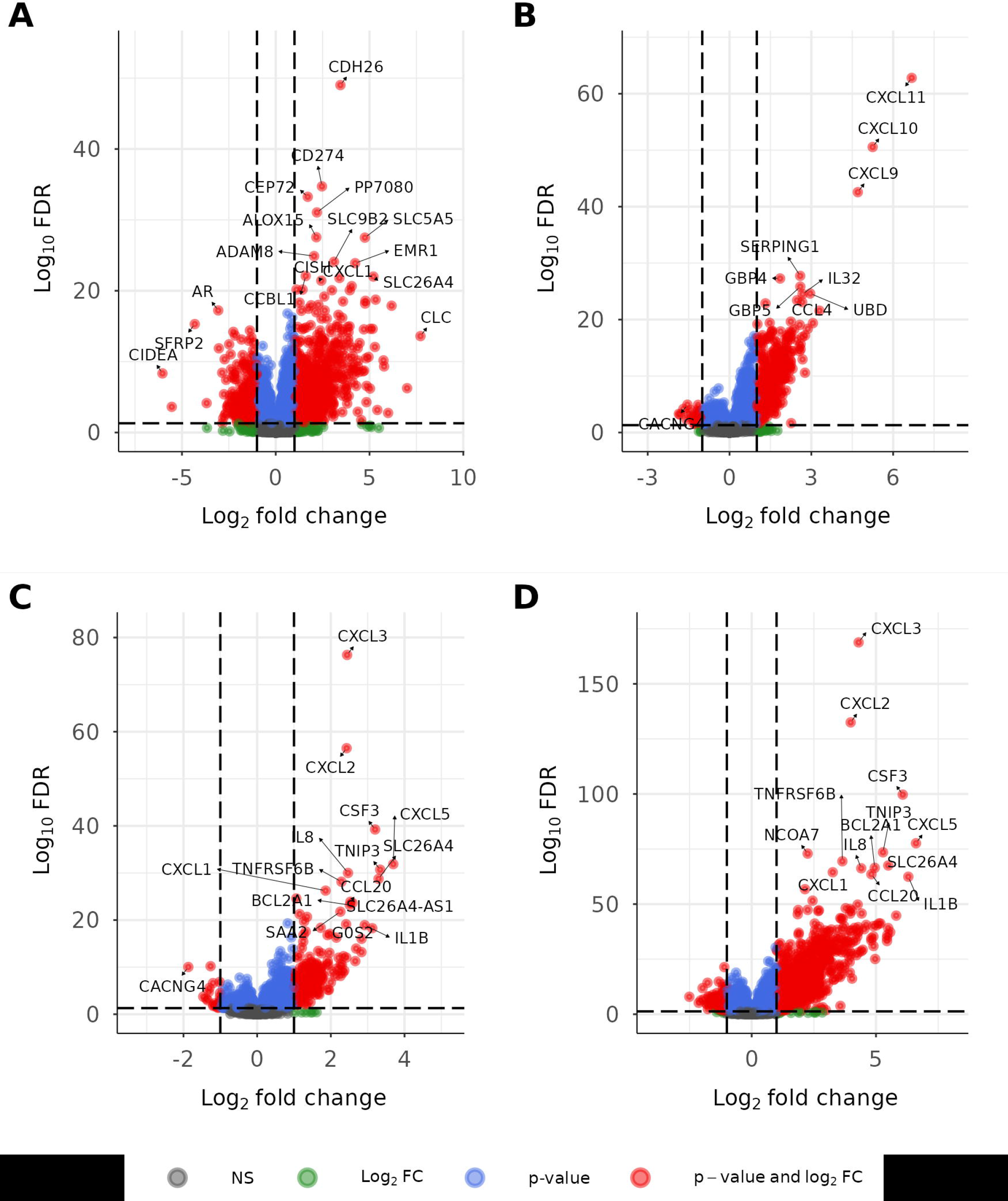
Differential analysis of gene expression for each asthma endotype in SOL-Asthma, with T2^LOW^/T1^LOW^/T17^LOW^ asthma as the comparison group. <u>Footnote</u>: Panel A shows differentially expressed genes (DEGs) in cases with T2^HIGH^ asthma. Panel B shows DEGs in cases with T1^HIGH^ asthma. Panel C shows DEGs in cases with T17^HIGH^ asthma. Panel D shows DEGs in cases with T1^HIGH^/T17^HIGH^ asthma. Volcano plots show the –log10 false discovery rate-adjusted (FDR) P value (FDR-P) vs log2 fold change in expression level. Genes with low FDR-P values, corresponding to differential effects, appear at the top. Log2 fold change refers to the logarithm (base 2) of the fold change in expression between 2 groups, where fold change is the ratio of the expression level of a gene in each endotype group to the expression level in the comparison group. Red = |log2 fold change|>1 and FDR-adjusted P value <0.01. Blue = FDR <0.01 but fold change below threshold. Green = |log2 fold change|>1 but FDR-adjusted P value below significance threshold.

Using the same approach as in SOL-Asthma, we identified five nasal transcriptomic profiles in CAAPA cases: T2^HIGH^(n=25 [14.4%]), T1^HIGH^ (n=47 [27%]), T17^HIGH^ (n=23 [13.2%]),T1^HIGH^/T17^HIGH^ (n=31 [17.8%]), and T2^LOW^/T1^LOW^/T17^LOW^ (n=48 [27.6%]) (**Figure 2, Panel B**). As in SOL-Asthma, k-means clustering and GMM classifications showed good concordance (**eFigure 5**).

**eTable 8** shows the characteristics of CAAPA participants by asthma status and endotype. Compared with controls, all cases had lower %predicted pre-BD FEV1 and pre-BD FEV1/FVC and cases with: 1) T2^LOW^/T1^LOW^/T17^LOW^asthma were older; 2) T1^HIGH^/T17^HIGH^ and T1^HIGH^ asthma had higher BMI; 3) T2^HIGH^ and T17^HIGH^ asthma were more likely to live outside the U.S.; and 4) T1^HIGH^/T17^HIGH^ asthma were more likely to live in the U.S. In a multivariable analysis, age was associated with lower odds of T17^HIGH^ asthma; living outside the U.S. was associated with increased odds of T2^HIGH^and T17^HIGH^asthma but lower odds of T1^HIGH^/T17^HIGH^ and T2^LOW^/T1^LOW^/T17^LOW^ asthma; %predicted pre-BD FEV1 was inversely associated with T1^HIGH^/T17^HIGH^ and T17^HIGH^ asthma; and pre-BD FEV1/FVC was inversely associated with all endotypes (**eTable 9**).

**eFigures 7-8** show the results of the analyses of DEGs and pathway enrichment in CAAPA. In T2^HIGH^ asthma, 129 (14.5%) of 892 upregulated DEGs in SOL-Asthma were replicated in CAAPA, with genes such as *CLC*, *CLCA1*, and *CCR3* upregulated in both cohorts. In T1^HIGH^ asthma, 274 (70.8%) of 387 DEGs in SOL-Asthma were replicated in CAAPA, including *CXCL9*, *CXCL10*, and *CXCL11*. In T17^HIGH^ asthma, 137 (65.6%) of 209 DEGs in SOL were replicated in CAAPA, with consistent upregulation of genes such as *CXCL5*, *CSF3*, *IL1B*, *DEFB4A*, *CCL20*, and *TREM1*. In T1^HIGH^/T17^HIGH^ asthma, 528 (54.0%) of 977 DEGs in SOL-Asthma were replicated in CAAPA, with *CXCL5*, *IL1B*, *CSF3*, *FPR1*, and *FPR2* upregulated in both cohorts.

## DISCUSSION

A previous study of gene networks for T2 and T1 pathways in sputum from 347 adults with asthma by Fahy et al identified four transcriptomic profiles: T2-high/T1-low (26.9%), T1-high/T2-low (13.5%), T2-high/T1-high ( 8.5%) and T2-low/T1-low (51.1%).^5^ A baseline T1-high profile was linked to SAEs, subclinical viral carriage, corticosteroid resistance, and persistence of this profile for 3 years, supporting T1-high asthma as an endotype. That study included mostly non-Hispanic Whites (66.3%), with smaller representation of non-Hispanic Blacks (22.8%) and Hispanics (3.5%).

The current study identified five transcriptomic profiles through expression of T2, T1 and T17 “signature genes” in nasal samples from Puerto Rican and Dominican adults, with replication of these profiles in a cohort of adults of African ancestry. Consistent with our prior findings in predominantly Puerto Rican and non-Hispanic Black youths,^7^ T2^HIGH^ asthma was less common than T2^LOW^ asthma endotypes.

Among U.S. adults, women and Puerto Ricans are disproportionately affected with asthma, and Puerto Ricans may be at increased risk of asthma-COPD overlap.^3,26,27^ In SOL-Asthma, Puerto Rican background was associated with T17^HIGH^ asthma and female sex was linked to T1^HIGH^, T1^HIGH^/T17^HIGH^and T2^LOW^/T1^LOW^/T17^LOW^asthma, while COPD was associated with T17^HIGH^, T1^HIGH^/T17^HIGH^ and T2^LOW^/T1^LOW^/T17^LOW^ asthma. Based on our prior findings in youths, Puerto Rican adults may have greater risk of T17^HIGH^ asthma than Dominican adults due to factors including different frequencies of risk polymorphisms, second-hand smoke exposure in childhood, dietary patterns, obesity, and violence-related distress.^28–31^ Similarly, sex hormones may underlie sex-specific findings for asthma endotypes, and this should be examined in future studies.

In SOL-Asthma, T1^HIGH^ asthma was strongly associated with SAEs in cross-sectional and prospective analyses, and T1^HIGH^, T1^HIGH^/T17^HIGH^and T2^LOW^/T1^LOW^/T17^LOW^asthma were retrospectively linked to FEV1/FVC decline over 11.4 to 16.7 years. Our findings suggest that adults with T1^HIGH^ asthma may be “exacerbation prone”, perhaps through enhanced susceptibility to viral infections.^5^ Nasal samples were collected ≥4 weeks after self-reported upper respiratory illnesses, but we did not assess subclinical viral carriage, which was previously correlated with T1^HIGH^ asthma.^5^ Fahy et al did not assess T17 pathways, and misclassification of T17^HIGH^ asthma may partly explain their findings of corticosteroid resistance in T1^HIGH^ asthma.

Despite divergent study designs and geographic locations, we replicated nasal transcriptomic profiles across studies, and there was substantial concordance in DEGs and enriched pathways between SOL-Asthma and CAAPA. However, we could not replicate most findings for potential risk factors in SOL-Asthma due to the smaller sample size and exclusion criteria in CAAPA. Interestingly, Puerto Rican background was associated with T17^HIGH^ and T1^HIGH^ and asthma in SOL-Asthma while living outside the U.S. was associated with T17^HIGH^ and T2^HIGH^ asthma in CAAPA.

### Limitations

First, endotypes may vary across time and we have a single assessment. However, T1^HIGH^ and (particularly) T2^HIGH^ asthma were previously shown to be more likely to persist over three years^5^. Second, corticosteroids could alter gene expression. However, corticosteroid use is unlikely to explain our results, as we accounted for corticosteroid use in SOL-Asthma and our findings are consistent with those in minority youths unexposed to corticosteroids.^9^ Third, we show similar findings in analyses excluding participants who smoked or had physician-diagnosed COPD, but residual misclassification of asthma is possible in the absence of imaging. Fourth, we had limited statistical power for analysis of low-frequency endotypes (e.g., T2^HIGH^ asthma) or to test for interactions between endotypes and covariates such as birthplace. Lastly, the mixed cellularity of nasal samples could lead to immune cell-derived transcriptomic signals, though we previously showed predominance of nasal epithelial signals (known to correlate with those from bronchial epithelium).^14^

## CONCLUSION

Five nasal transcriptomic profiles were identified in a study of Puerto Rican and Dominican adults and replicated in a cohort of adults of African ancestry. Such profiles likely represent true asthma endotypes, since they were associated with distinct variables and outcomes. Our findings support further studies of nasal transcriptomics as a non-invasive approach to better understand asthma pathogenesis and develop much needed therapeutic approaches for T2-low asthma endotypes.

## Data Availability

All data produced in the present study are available upon reasonable request to the authors

## ACKNOWLEDGEMENTS

<u>Funding and role of funder</u>

The Study of Latinos (SOL)-Asthma Study was funded by grant HL152475 from the U.S. National Institutes of Health (NIH). Dr Yang is supported by a T32 Training Grant (HL129949) from the NIH. The Hispanic Community Health Study/Study of Latinos is a collaborative study supported by contracts from the National Heart, Lung, and Blood Institute (NHLBI) to the University of North Carolina (HHSN268201300001I / N01-HC-65233), University of Miami (HHSN268201300004I / N01-HC-65234), Albert Einstein College of Medicine (HHSN268201300002I / N01-HC-65235), University of Illinois at Chicago (HHSN268201300003I / N01-HC-65236 Northwestern Univ), and San Diego State University (HHSN268201300005I / N01-HC-65237). The following Institutes/Centers/Offices have contributed to the HCHS/SOL through a transfer of funds to the NHLBI: National Institute on Minority Health and Health Disparities, National Institute on Deafness and Other Communication Disorders, National Institute of Dental and Craniofacial Research, National Institute of Diabetes and Digestive and Kidney Diseases, National Institute of Neurological Disorders and Stroke, NIH Institution-Office of Dietary Supplements. The Consortium on Asthma among African Ancestry Populations in the Americas (CAAPA) was funded by NIH grant HL104608. This current analysis in CAAPA was supported in part by the Intramural Research Program of NIH. The contributions of the NIH authors were made as part of their official duties as NIH federal employees, are in compliance with agency policy requirements, and are considered Works of the U.S. Government. However, the findings and conclusions presented in this paper are those of the authors and do not necessarily reflect the views of the NIH or the U.S. Department of Health and Human Services.

## Access to Data and Data Analysis

Dr. Sheng, Dr. Han, Dr. Chen and Dr. Celedón had full access to all data in the study and take responsibility for the integrity of the data and the accuracy of the data analysis.

## Data Sharing

RNA-Seq data from SOL-Asthma will be deposited in a public database upon acceptance of the manuscript for publication. RNA-Seq data from CAAPA were accessed from the previously deposited data in GEO (<u>GSE240567</u>).

## REFERENCES

1. Asher MI, García-Marcos L, Pearce NE, Strachan DP. Trends in worldwide asthma prevalence. European Respiratory Journal. 2020;56(6):2002094. doi:10.1183/13993003.02094-2020

2. Porsbjerg C, Melén E, Lehtimäki L, Shaw D. Asthma. The Lancet. 2023;401(10379):858–873. doi:10.1016/S0140-6736(22)02125-0

3. Barr RG, Avilés-Santa L, Davis SM, et al. Pulmonary Disease and Age at Immigration among Hispanics. Results from the Hispanic Community Health Study/Study of Latinos. American Journal of Respiratory and Critical Care Medicine. 2016;193(4):386–395. doi:10.1164/rccm.201506-1211OC

4. Nazario S, Telón-Sosa B, Arroyo-Cruz A, Guo K. Migration, educational, and asthma mortality disparities in the United States. The Journal of Allergy and Clinical Immunology: In Practice. 2025;13(7):1852–1854.e2. doi:10.1016/j.jaip.2025.03.027

5. Fahy JV, Jackson ND, Sajuthi SP, et al. Type 1 Immune Responses Related to Viral Infection Influence Corticosteroid Response in Asthma. American Journal of Respiratory and Critical Care Medicine. 2025;211(2):194–204. doi:10.1164/rccm.202402-0403OC

6. Gauthier M, Kale SL, Ray A. T1-T2 Interplay in the Complex Immune Landscape of Severe Asthma. Immunological Reviews. 2025/03/01 2025;330(1):e70011. 10.1111/imr.70011

7. Raundhal M, Morse C, Khare A, et al. High IFN-γ and low SLPI mark severe asthma in mice and humans. The Journal of Clinical Investigation. 06/29/ 2015;125(8):3037-3050. doi:10.1172/JCI80911

8. Woodruff PG, Modrek B, Choy DF, et al. T-helper Type 2–driven Inflammation Defines Major Subphenotypes of Asthma. American Journal of Respiratory and Critical Care Medicine. 2009;180(5):388–395. doi:10.1164/rccm.200903-0392OC

9. Yue M, Gaietto K, Han YY, et al. Transcriptomic Profiles in Nasal Epithelium and Asthma Endotypes in Youth. JAMA. 2025;333(4):307–318. doi:10.1001/jama.2024.22684

10. Sorlie PD, Avilés-Santa LM, Wassertheil-Smoller S, et al. Design and Implementation of the Hispanic Community Health Study/Study of Latinos. Annals of Epidemiology. 2010/08/01/ 2010;20(8):629–641. 10.1016/j.annepidem.2010.03.015

11. LaVange LM, Kalsbeek WD, Sorlie PD, et al. Sample Design and Cohort Selection in the Hispanic Community Health Study/Study of Latinos. Annals of Epidemiology. 2010/08/01/ 2010;20(8):642-649. 10.1016/j.annepidem.2010.05.006

12. Burkart KM, Sofer T, London SJ, et al. A Genome-Wide Association Study in Hispanics/Latinos Identifies Novel Signals for Lung Function. The Hispanic Community Health Study/Study of Latinos. American Journal of Respiratory and Critical Care Medicine. 2018;198(2):208–219. doi:10.1164/rccm.201707-1493OC

13. Nathan RA, Sorkness CA, Kosinski M, et al. Development of the asthma control test: A survey for assessing asthma control. Journal of Allergy and Clinical Immunology. 2004;113(1):59–65. doi:10.1016/j.jaci.2003.09.008

14. Forno E, Wang T, Qi C, et al. DNA methylation in nasal epithelium, atopy, and atopic asthma in children: a genome-wide study. The Lancet Respiratory Medicine. 2019;7(4):336–346. doi:10.1016/S2213-2600(18)30466-1

15. Miller MR, Hankinson J, Brusasco V, et al. Standardisation of spirometry. European Respiratory Journal. 26(2):319–338. doi:10.1183/09031936.05.00034805

16. Graham BL, Steenbruggen I, Miller MR, et al. Standardization of Spirometry 2019 Update. An Official American Thoracic Society and European Respiratory Society Technical Statement. American Journal of Respiratory and Critical Care Medicine. 2019;200(8):e70–e88. doi:10.1164/rccm.201908-1590ST

17. Szczesny B, Boorgula MP, Chavan S, et al. Multi-omics in nasal epithelium reveals three axes of dysregulation for asthma risk in the African Diaspora populations. Nature Communications. 2024/05/28 2024;15(1):4546. doi:10.1038/s41467-024-48507-7

18. Ewels PA, Peltzer A, Fillinger S, et al. The nf-core framework for community-curated bioinformatics pipelines. Nature Biotechnology. 2020/03/01 2020;38(3):276–278. doi:10.1038/s41587-020-0439-x

19. Woodruff PG, Boushey HA, Dolganov GM, et al. Genome-wide profiling identifies epithelial cell genes associated with asthma and with treatment response to corticosteroids. Proceedings of the National Academy of Sciences. 2007;104(40):15858–15863. doi:doi:10.1073/pnas.0707413104

20. Choy DF, Hart KM, Borthwick LA, et al. T_H_2 and T_H_17 inflammatory pathways are reciprocally regulated in asthma. Science Translational Medicine. 2015;7(301):301ra129–301ra129. doi:doi:10.1126/scitranslmed.aab3142

21. Charrad M, Ghazzali N, Boiteau V, Niknafs A. NbClust: An R Package for Determining the Relevant Number of Clusters in a Data Set. Journal of Statistical Software. 11/03 2014;61(6):1-36. doi:10.18637/jss.v061.i06

22. Lumley T. Analysis of Complex Survey Samples. Journal of Statistical Software. 04/15 2004;9(8):1–19. doi:10.18637/jss.v009.i08

23. Bowerman C, Bhakta NR, Brazzale D, et al. A Race-neutral Approach to the Interpretation of Lung Function Measurements. American Journal of Respiratory and Critical Care Medicine. 2023;207(6):768–774. doi:10.1164/rccm.202205-0963OC

24. Love MI, Huber W, Anders S. Moderated estimation of fold change and dispersion for RNA-seq data with DESeq2. Genome Biology. 2014/12/05 2014;15(12):550. doi:10.1186/s13059-014-0550-8

25. Wu T, Hu E, Xu S, et al. clusterProfiler 4.0: A universal enrichment tool for interpreting omics data. The Innovation. 2021/08/28/ 2021;2(3):100141. 10.1016/j.xinn.2021.100141

26. Han Y-Y, Forno E, Celedón JC. Sex Steroid Hormones and Asthma in a Nationwide Study of U.S. Adults. American Journal of Respiratory and Critical Care Medicine. 2020;201(2):158–166. doi:10.1164/rccm.201905-0996OC

27. Han Y-Y, Forno E, Canino G, Celedón JC. Psychosocial risk factors and asthma among adults in Puerto Rico. Journal of Asthma. 2019/06/03 2019;56(6):653–661. doi:10.1080/02770903.2018.1474366

28. Joe A, Rosser FJ, Gaietto K, Yue M, Chen W, Han Y-Y, Celedón JC. Dietary Patterns and Asthma Endotypes in Puerto Rican Youth. CHEST. doi:10.1016/j.chest.2026.05.012

29. Han Y-Y, Yue M, Pereira K, Rosser FJ, Gaietto K, Chen W, Celedón JC. Persistent exposure to second-hand smoke and asthma endotypes in a study of Puerto Rican youth. Annals of Allergy, Asthma & Immunology. 2026/06/01/ 2026;136(6):665–670.e2. 10.1016/j.anai.2026.03.011

30. Adkins AD, Han Y-Y, Yue M, et al. Overweight, obesity, adiposity measures, and asthma endotypes in Puerto Rican youth. American Journal of Respiratory and Critical Care Medicine. 2026;212(4):830–833. doi:10.1093/ajrccm/aamaf131

31. Yue M, Gaietto K, Xu Z, et al. Violence-Related Distress, Nasal Epithelial Gene Expression, and T17-High Asthma in Youth. Pediatric Pulmonology. 2026/02/01 2026;61(2):e71486. 10.1002/ppul.71486

