## Supplementary material for "Nasal transcriptomics characterize T-helper (T)2-low asthma endotypes in a study of Puerto Rican and Dominican adults living in the U.S.": Online Supplement

### **eMethods**

#### ***Study of Latinos (SOL)-Asthma Study***

At the baseline in-person visit for the SOL Asthma Study, all participants completed a protocol including questionnaires on demographics and respiratory health, spirometry, and collection of nasal samples. Participants with asthma (cases) also completed the Asthma Control Test (ACT) questionnaire.^1^ Spirometry was conducted with an EasyOne (NDD Medical Technologies, Andover, MA, USA) spirometer, following American Thoracic Society/European Respiratory Society recommendations.^2^ Spirometry was repeated 15 minutes after administration of two puffs of inhaled albuterol. Subjects were asked to withhold their inhaled bronchodilators (BD) -when possible- before spirometry testing (e.g., 4-6 hours and 24 hours for short- and long-acting ß2-agonists, respectively, and 36 hours for ultra-long-acting ß2-agonists). All spirometry tests were graded based on the within- and between-maneuver acceptability and repeatability criteria. Grades of “A”, “B”, or “C” were considered of good quality and included in the analysis. One year after the baseline in-person visit, we conducted a phone call in participants with asthma (cases). This call included administration of the ACT questionnaire and a short questionnaire on emergency department visits, hospitalizations, use of inhaled corticosteroids (ICS), and use of oral corticosteroids (OCS) for asthma since the baseline visit.

We conducted a prospective analysis of indicators of asthma severity or control in cases. In this analysis, our first outcome of interest was at least one severe asthma exacerbation (defined as at least one ED visit or at least one hospitalization for asthma) between the baseline visit and the one-year follow-up phone call. In addition to the covariates included in our main analysis of asthma endotypes, this analysis was adjusted for ICS use, categorized as: 1) “Both visits” if ICS use was reported at both the baseline visit and the one-year follow-up visit (by phone call), 2) “Either visit”, if ICS use was reported at one visit but not the other, and 3) “Never” if ICS use was reported at neither visit. Our second outcome of interest for the prospective analysis was a decline (negative change) of three or more points in the ACT score (considered clinically meaningful) between the baseline and follow-up visits. This analysis was adjusted for the same covariates as that of ED visits or hospitalizations for asthma, except that we adjusted for baseline ACT score instead of baseline lung function measures.

#### ***Consortium on Asthma among African ancestry Populations in the Americas (CAAPA)***

Detailed information about subject recruitment and study protocol, data collection, RNA sequencing, ancestry deconvolution (ADMIXTURE), and principal components analysis in CAAPA were previously published.^3^

Eligibility criteria included residence in four U.S. sites (Denver, Baltimore, Washington DC, and Chicago) and three non-US sites (Barbados, Nigeria, and Salvador in Brazil); self-identification as African, African American, African Caribbean, African Brazilian, or African-Other; and ability to provide written informed consent. Asthma was defined as ever asthma confirmed by a physician, with affirmative responses to both 'Have you ever had asthma?' and 'Was it confirmed by a doctor?'. Current asthma was further defined as a Composite Asthma Severity Index (CASI) score ≥ 1 at the time of enrollment.^4,5^ Controls never had asthma. RNA -Seq was conducted in RNA extracted from nasal samples of participants with (cases, *n*=253) and without (controls, *n*=283) current asthma.

In contrast to SOL-Asthma, adults who had been diagnosed with COPD were excluded from CAAPA, each recruitment site was balanced for age and sex between cases and controls, and only one participant was current smoker. Additional parameters of interest used in this replication assessment (i.e. medications and hospitalizations) were taken from the CASI which takes into consideration medication use and the corresponding treatment level in determining asthma severity.^3^ Atopy was defined as total serum IgE (tIgE)>100 kU/L defined based on Wong et al.^6^ and/or multi-allergen ImmunoCAP phadiatop specific serum IgE (sIgE) ≥ 0.36 PAU/L per assay protocol recommendations.

Following the same procedure as in SOL-Asthma, we used mclust (version 6.1.1) to fit Gaussian Mixture Models in a sensitivity analysis, using the same 13 “signature genes” as those included in the analysis using a K-means clustering approach.^7^ We used dplyr (version 1.1.4) and plyr (version 1.8.9) for data processing.

As in SOL-Asthma, we analyzed DEGs between each endotype and T2^LOW^/T1^LOW^/T17^LOW^ asthma adjusting for sex, age, study site, RNA integrity number, CG content, library batch, and two genetic PCs. We then conducted a pathway enrichment analysis for upregulated genes.

#### ***Weighted gene co-expression network analysis (WCGNA) in SOL-Asthma***

We conducted a consensus Weighted Gene Co-expression Network Analysis (WGCNA) to investigate the gene modules associated with asthma endotypes in SOL-Asthma, excluding the 13 “signature” genes used in the main analyses.^8^ First, we performed soft-thresholding power network topology analysis separately for each cohort and then determined a suitable power value. We then constructed a network based on the adjacency matrix, which was then transformed into a topological overlap matrix (TOM) to estimate the distance between each pair of genes. Next, we employed hierarchical clustering with average and dynamic methods to construct a cluster tree and classify genes into different modules. To assess the associations between modules and asthma endotypes, we calculated the correlations of the consensus module eigengene (ME) with the endotypes for each cohort. Candidate gene modules were defined as those with correlations with endotypes greater than 0.15.

#### ***Refining gene expression profiles for T2-high, T1-high, and T17-high asthma***

Based on the endotype-specific gene modules we identified using WGCNA and genes that were significant (log FC>1 and FDR<0.05) in the analysis of differentially expressed genes, we built a new signature gene panel. Using the T2, T1, and T17 candidate genes identified, we built Lasso binary classification prediction models for T2-high, T1-high, and T17-high asthma respectively using individuals in the two study cohorts with a consensus profile.

### **eResults**

#### ***Refining gene expression profiles for T2^HIGH^, T1^HIGH^, and T17^HIGH^ asthma***

Using the gene modules identified in WGCNA and genes that were significant in the analysis of differential expression, we identified candidate genes for T2-high (n=50), T1-high (n=499), and T17-high (n=345) asthma. Using Lasso binary classification prediction models, we identified 3 genes for the refined T2 panel; 23 genes for the refined T1 panel and 9 genes for the refined T17 panel. Heat maps of expression levels for the gene panels in SOL-Asthma are shown in eFigure 7 and demonstrate a similar distribution for the five endotypes as that using the 13 “signature genes”.

T1 immunity in asthma was previously shown to be characterized by the accumulation of IFN-γ-producing tissue-resident memory T cells (TRM) in the airways, with CD8+ and CD4+ TRM cells as key contributors to T1-driven airway inflammation and disease severity.^9,10^ Consistent with this, LASSO analysis of differentially expressed genes in T1^HIGH^ asthma identified upregulation of TRM markers, including *ZNF683* and *CXCR6.*^11,12^

#### ***Pathway enrichment analysis***

Enriched canonical pathways for upregulated DEGs for each asthma endotype (at FDR-P <0.01) were identified by ingenuity pathway analysis in both SOL-Asthma and CAAPA.

#### **SOL-Asthma**

In T2^HIGH^ asthma, there were 645 significantly enriched pathways, with those specific to type 2 inflammation including eosinophil migration, mast cell activation and degranulation, and IL-4 production. In T1^HIGH^ asthma, 652 pathways were enriched, with the strongest enrichment for type II interferon production, leukocyte-mediated cytotoxicity, T cell mediated immunity, and natural killer cell mediated immunity. In T17^HIGH^ asthma, 452 pathways were enriched, including neutrophil chemotaxis, granulocyte migration, acute-phase response, and antimicrobial humoral response. In T1^HIGH^/T17^HIGH^ asthma, there was concurrent enrichment of both T1-related (e.g., type II interferon production and leukocyte-mediated cytotoxicity), and T17-related (e.g., neutrophil migration and granulocyte chemotaxis) pathways, reflecting the dual T1/T17 inflammatory pathways for this endotype.

**CAAPA**

Given small sample size, only eight pathways were significantly enriched in T2^HIGH^ asthma in CAAPA. Of these, collagen-containing extracellular matrix and histamine transport were consistently enriched in both cohorts, consistent with T2-driven airway remodeling and mast cell activation. In T1^HIGH^ asthma, 518 (79.4%) of 652 pathways enriched in SOL-Asthma were replicated in CAAPA, including type II interferon production, leukocyte mediated cytotoxicity, T cell mediated immunity, and natural killer cell mediated immunity. In T17^HIGH^ asthma, 299 (66.2%) of 452 pathways enriched in SOL-Asthma were replicated in CAAPA, including neutrophil migration, granulocyte migration, cell chemotaxis, and activation of innate immune response. Of 1,149 pathways enriched for T1^HIGH^/T17^HIGH^ asthma in SOL-Asthma, 753 (65.5%) were replicated in CAAPA, with concurrent enrichment of T1-related (e.g., type II interferon production, leukocyte mediated cytotoxicity, and CD8+ alpha-beta T cell activation) and T17-related (e.g., neutrophil migration, granulocyte chemotaxis, and interleukin-17 production) pathways.

**REFERENCES**

1.	Nathan RA, Sorkness CA, Kosinski M, et al. Development of the asthma control test: A survey for assessing asthma control. *Journal of Allergy and Clinical Immunology*. 2004;113(1):59-65. doi:10.1016/j.jaci.2003.09.008

2.	Miller MR, Hankinson J, Brusasco V, et al. Standardisation of spirometry. *European Respiratory Journal*. 26(2):319-338. doi:10.1183/09031936.05.00034805

3.	Szczesny B, Boorgula MP, Chavan S, et al. Multi-omics in nasal epithelium reveals three axes of dysregulation for asthma risk in the African Diaspora populations. *Nature Communications*. 2024/05/28 2024;15(1):4546. doi:10.1038/s41467-024-48507-7

4.	Krouse RZ, Sorkness CA, Wildfire JJ, et al. Minimally important differences and risk levels for the Composite Asthma Severity Index. *Journal of Allergy and Clinical Immunology*. 2017;139(3):1052-1055. doi:10.1016/j.jaci.2016.08.041

5.	Wildfire JJ, Gergen PJ, Sorkness CA, et al. Development and validation of the Composite Asthma Severity Index-an outcome measure for use in children and adolescents. *Journal of Allergy and Clinical Immunology*. 2012;129(3):694-701. doi:10.1016/j.jaci.2011.12.962

6.	Wong C-Y, Yeh K-W, Huang J-L, et al. Longitudinal analysis of total serum IgE levels with allergen sensitization and atopic diseases in early childhood. *Scientific Reports*. 2020/12/04 2020;10(1):21278. doi:10.1038/s41598-020-78272-8

7.	Scrucca L, Fop M, Murphy TB, Raftery AE. mclust 5: Clustering, Classification and Density Estimation Using Gaussian Finite Mixture Models. *R j*. Aug 2016;8(1):289-317.

8.	Langfelder P, Horvath S. WGCNA: an R package for weighted correlation network analysis. *BMC Bioinformatics*. 2008/12/29 2008;9(1):559. doi:10.1186/1471-2105-9-559

9.	Camiolo MJ, Zhou X, Oriss TB, et al. High-dimensional profiling clusters asthma severity by lymphoid and non-lymphoid status. *Cell Reports*. 2021;35(2)doi:10.1016/j.celrep.2021.108974

10.	Herrera-De La Mata S, Ramírez-Suástegui C, Mistry H, et al. Cytotoxic CD4+ tissue-resident memory T cells are associated with asthma severity. *Med*. 2023/12/08/ 2023;4(12):875-897.e8. doi:<https://doi.org/10.1016/j.medj.2023.09.003>

11.	Wein AN, McMaster SR, Takamura S, et al. CXCR6 regulates localization of tissue-resident memory CD8 T cells to the airways. *Journal of Experimental Medicine*. 2019;216(12):2748-2762. doi:10.1084/jem.20181308

12.	Mackay LK, Minnich M, Kragten NAM, et al. Hobit and Blimp1 instruct a universal transcriptional program of tissue residency in lymphocytes. *Science*. 2016;352(6284):459-463. doi:doi:10.1126/science.aad2035

### **eFigures**

#### **eFigure 1. Optimal number of clusters determined by NbClust majority rule.**

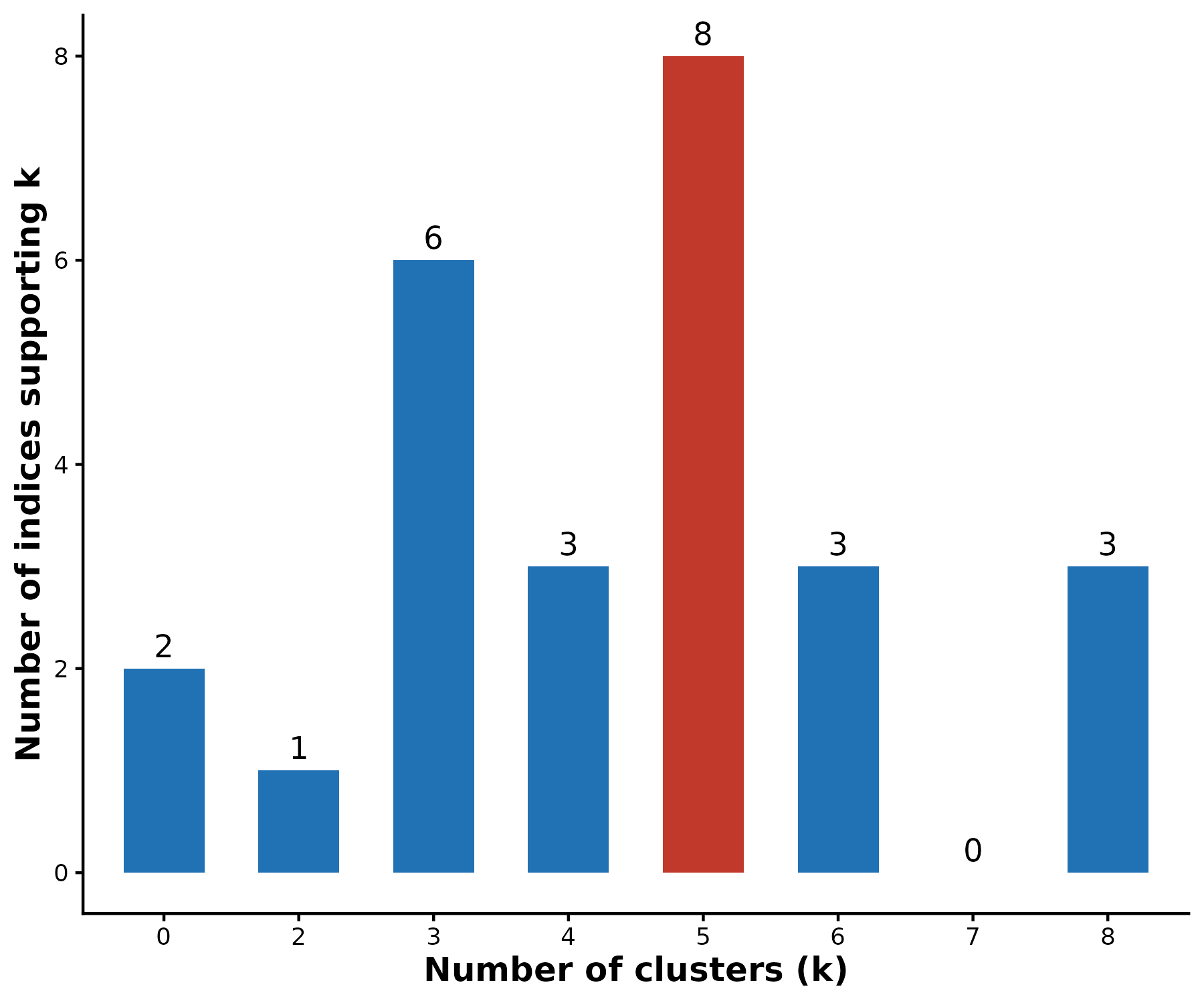

Because of high computational cost, four indices (Gap, Gamma, Gplus, and Tau) were not computed when *index* was set to "all". Hubert and Dindex (k=0) are graphical indices that do not provide a numeric optimal k, and Frey (k=2) returned a value outside the specified range (k=3–8). Of the 23 applicable indices, 8 supported k=5 as the optimal solution: Krzanowski-Lai (KL), Hartigan, Cubic Clustering Criterion (CCC), Scott, Marriot, Rubin, Davies-Bouldin (DB), and Ratkowsky, indicated by the red bar.

#### **eFigure 2. Heatmap of nasal transcriptomic profiles using K-means clustering and Gaussian Mixture Modeling approaches in SOL-Asthma (k=5).**

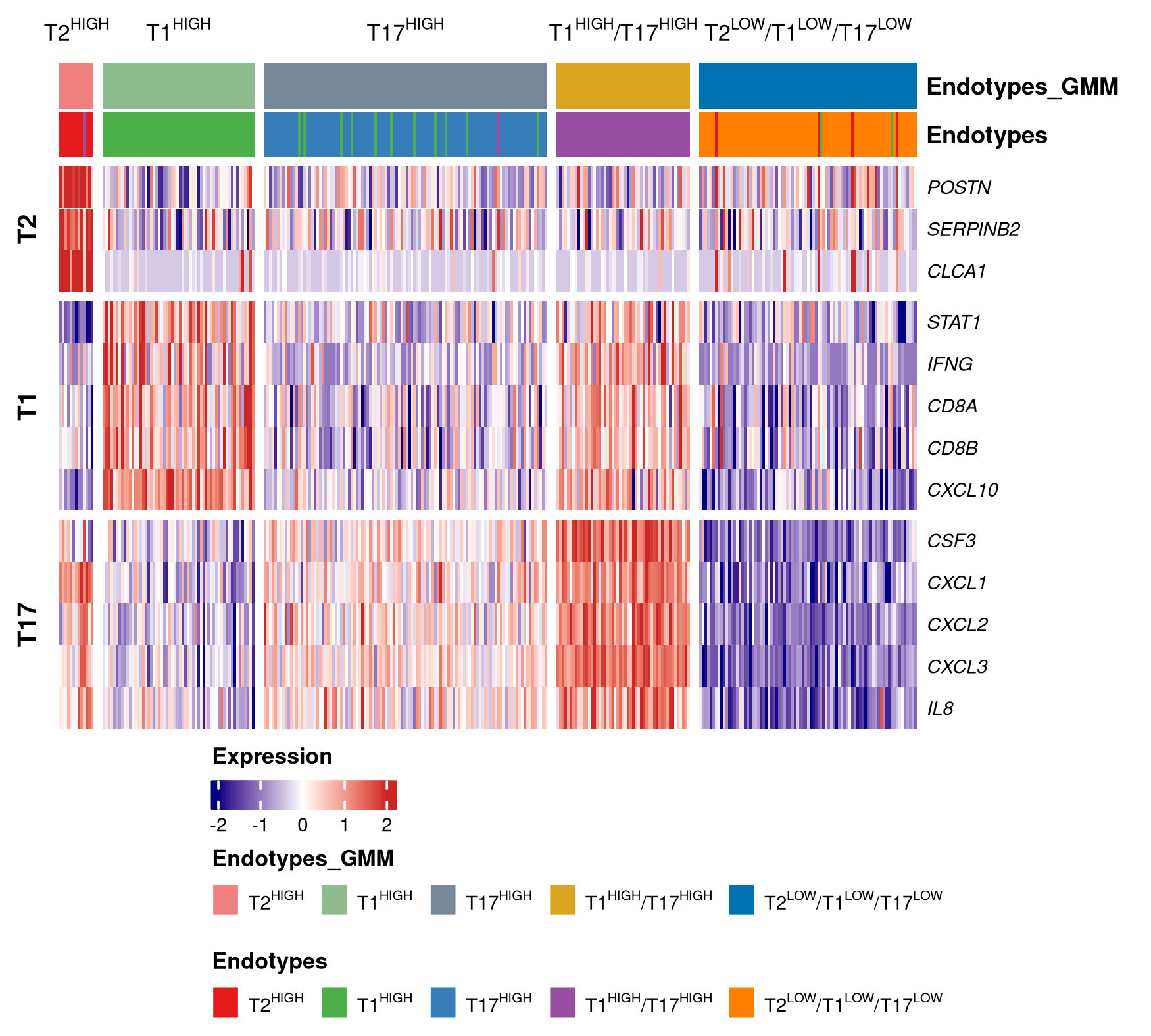

The five transcriptomic profiles by GMM were: T2^HIGH^ (n=13 [4.2%]), T1^HIGH^ (n=58 [18.5%]), T17^HIGH^ (n=108 [34.5%]), T1^HIGH^/T17^HIGH^ (n=51 [16.3%]), and T2^LOW^/T1^LOW^/T17^LOW^ (n=83 [22.5%]). 294 (93.9%) participants with asthma were assigned to the same endotype by both K-means clustering and GMM classification, and the Adjusted Rand Index (ARI) for the agreement between k-means and GMM classifications was 0.86.

#### **eFigure 3. Dotplot of the top 15 enriched pathways in SOL-Asthma, with T2^LOW^/T1^LOW^/T17^LOW^ asthma as the reference group.**

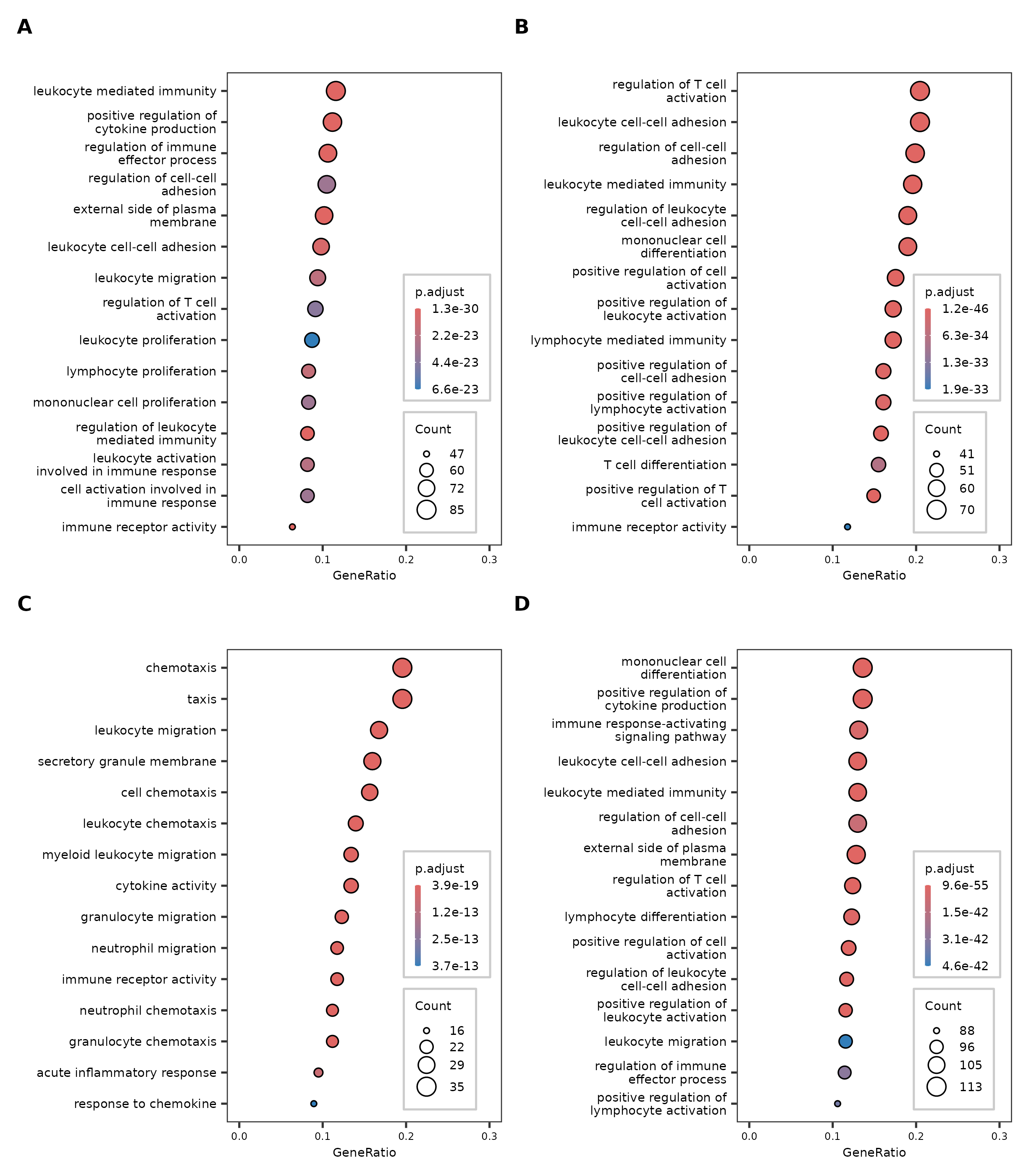

Panel **A** shows enriched pathways in cases with T2^HIGH^ asthma. Panel **B** shows enriched pathways in cases with T1^HIGH^ asthma. Panel **C** shows enriched pathways in cases with T17^HIGH^ asthma. Panel **D** shows enriched pathways in cases with T1^HIGH^/T17^HIGH^ asthma.

#### **eFigure 4. Heatmap generated using alternative genes for T2^HIGH^, T1^HIGH^, and T17^HIGH^ in SOL-Asthma (k=5).**

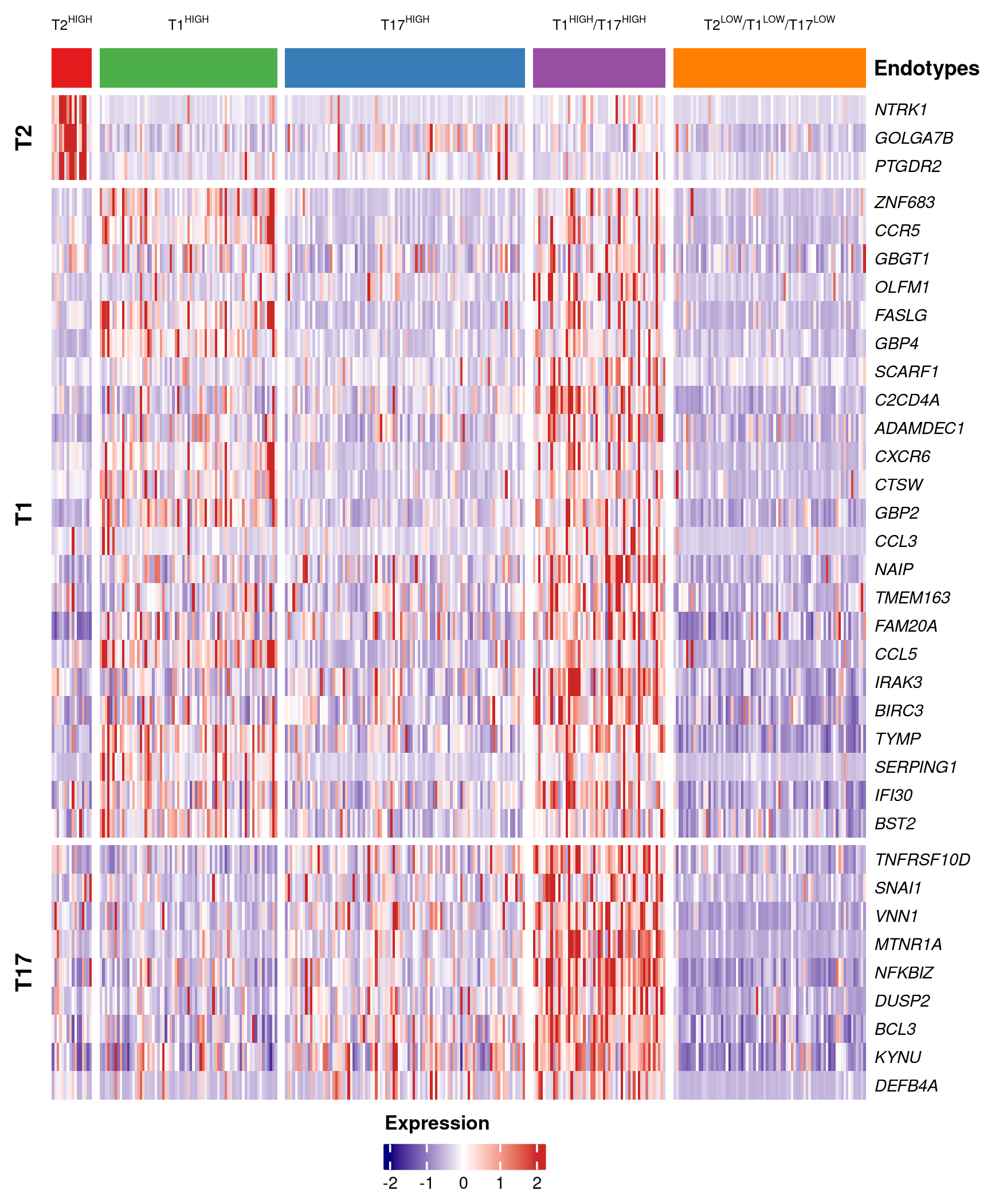

#### **eFigure 5. Heatmap of nasal transcriptomic profiles derived using a K-Means clustering approach and Gaussian Mixed Models in CAAPA (k=5).**

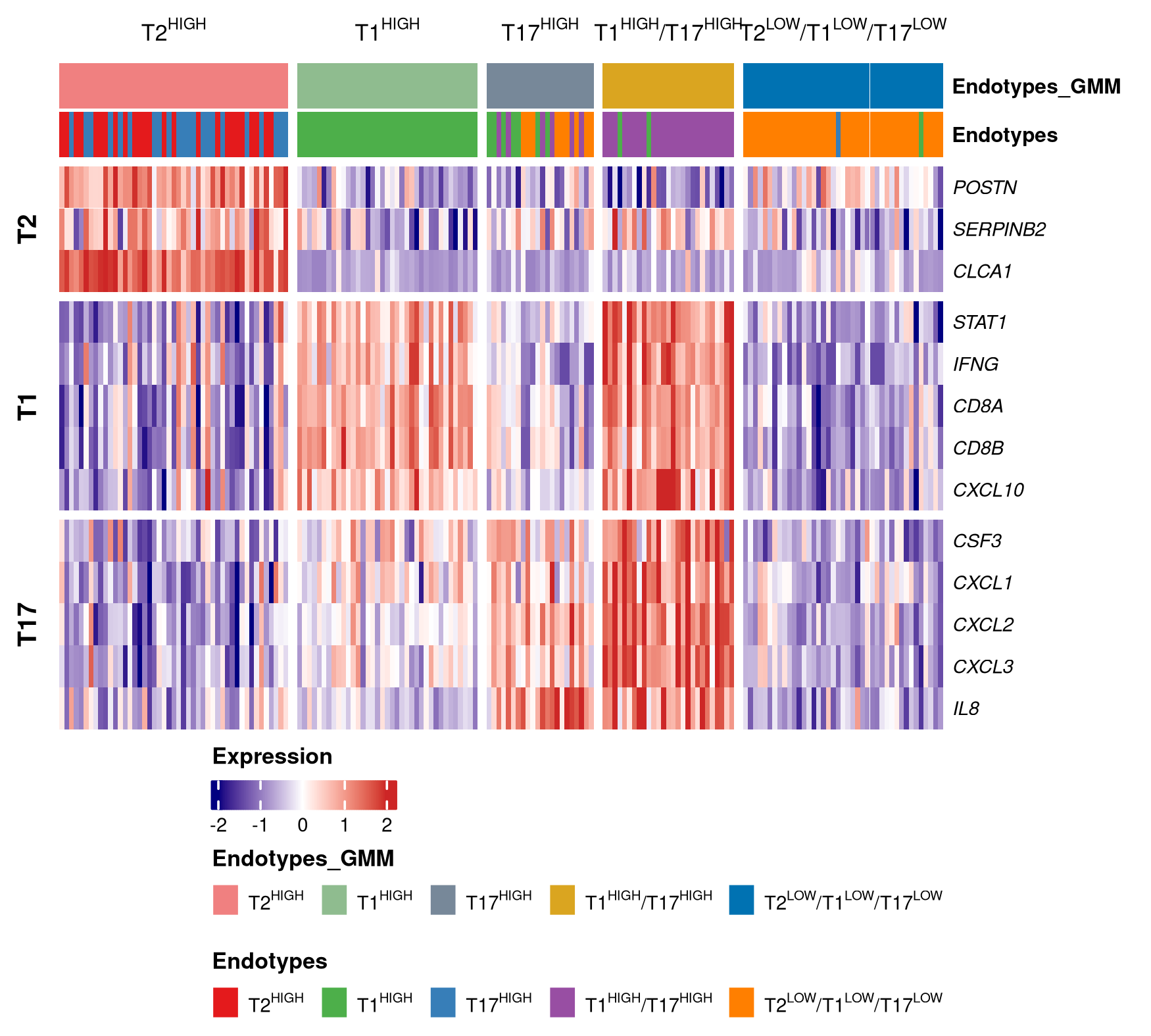

The five transcriptomic profiles by GMM were: T2^HIGH^ (n=47 [27.0%]), T1^HIGH^ (n=37 [21.3%]), T17^HIGH^ (n=22 [12.6%]), T1^HIGH^/T17^HIGH^ (n=27 [15.5%]), and T2^LOW^/T1^LOW^/T17^LOW^ (n=41 [23.6%]). 126 (72.4%) participants with asthma were assigned to the same endotype by both K-means clustering and GMM classification, with Adjusted Rand Index (ARI)=0.65.

#### **eFigure 6. Analysis of differential gene expression among cases in CAAPA, comparing each endotype to T2^LOW^/T1^LOW^/T17^LOW^ asthma.**

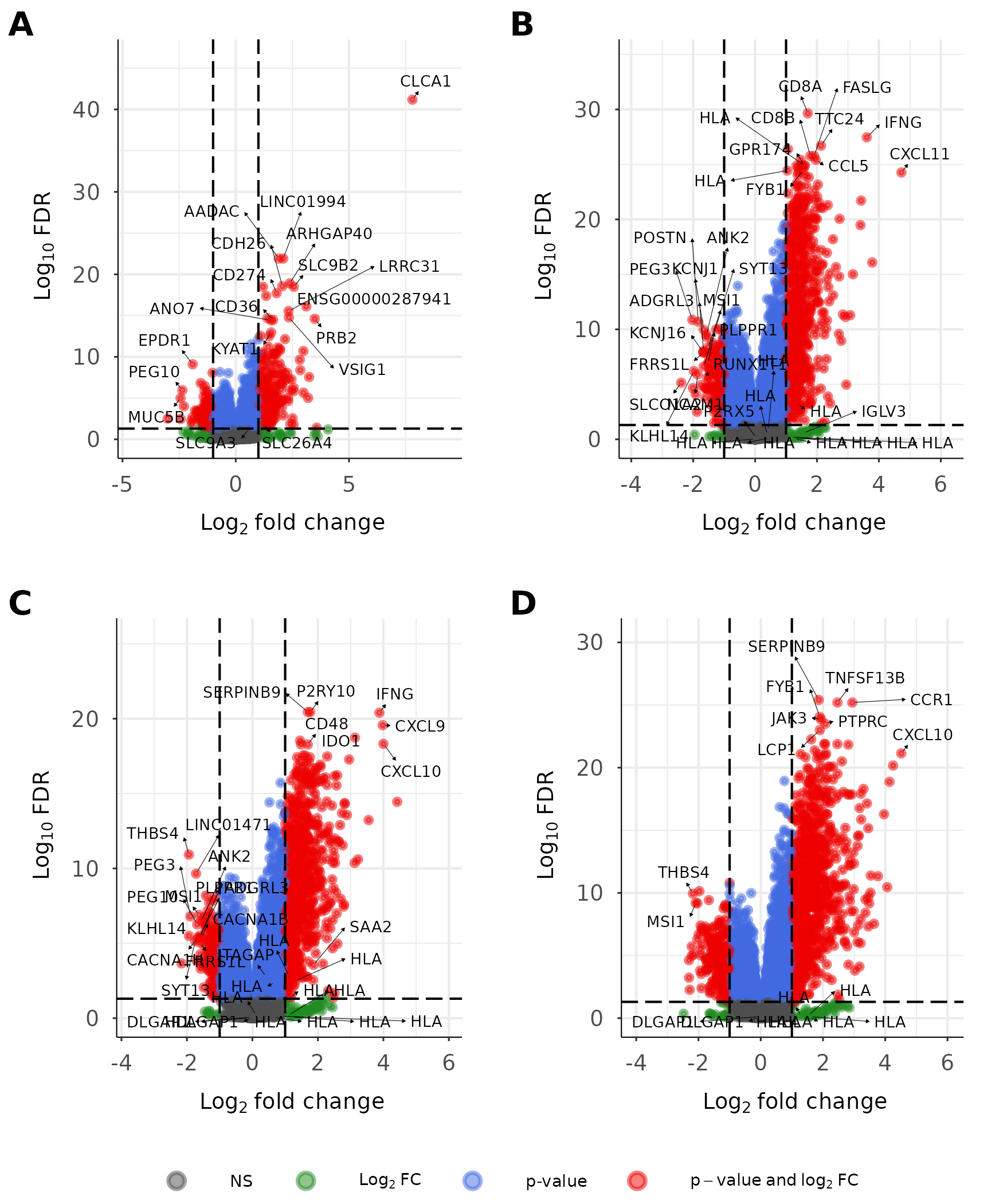

Panel **A** shows differentially expressed genes (DEGs) in cases with T2^HIGH^ asthma. Panel **B** shows DEGs in cases with T1^HIGH^ asthma. Panel **C** shows DEGs in cases with T17^HIGH^ asthma. Panel **D** shows DEGs in cases with T1^HIGH^/T17^HIGH^ asthma.

#### **eFigure 7. Dotplot of enriched pathways within each asthma endotype in CAAPA, with T2^LOW^/T1^LOW^/T17^LOW^ asthma as the reference group.**

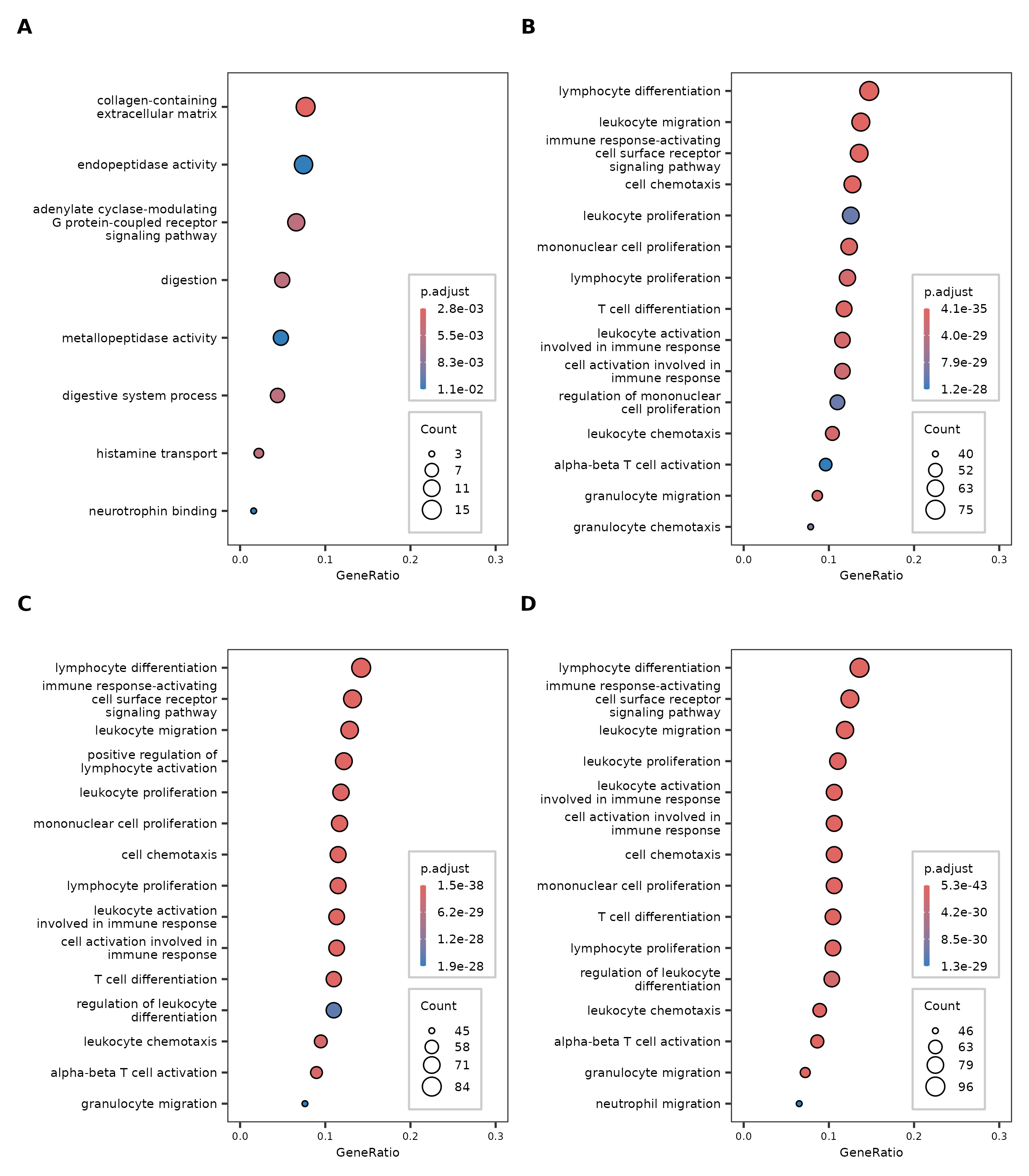

Panel **A** shows enriched pathways in cases with T2^HIGH^ asthma. Panel **B** shows enriched pathways in cases with T1^HIGH^ asthma. Panel **C** shows enriched pathways in cases with T17^HIGH^ asthma. Panel **D** shows enriched pathways in cases with T1^HIGH^/T17^HIGH^ asthma

### **eTables**

#### **eTable 1. Multinomial logistic regression analysis of asthma endotypes in SOL-Asthma (with controls as the reference outcome category), excluding participants who had physician-diagnosed COPD and/or were current smokers or former smokers with ≥10 pack-years of smoking**

|  | Asthma endotypes | | | | |
| --- | --- | --- | --- | --- | --- |
| Variables | **T2^HIGH^**  **(*n*=12)** | **T1^HIGH^**  **(*n*=46)** | **T17^HIGH^**  **(*n*=60)** | **T1^HIGH^/T17^HIGH^**  **(*n*=40)** | **T2^LOW^/T17^LOW^/T1^LOW^**  **(*n*=36)** |
|  | **Odds ratio (95% confidence interval)** | | | | |
| Age (years) | 0.94(0.85,1.03) | 1.00(0.96,1.03) | 1.00(0.96,1.03) | 0.96(0.93,1.00) | 1.05(1.00,1.11) |
| Female sex | 2.50(0.63,9.94) | 2.08(0.69,6.27) | 1.55(0.62,3.89) | **2.50(1.03,6.09)^†^** | 3.62(1.00,13.18) |
| Body mass index (kg/m^2^) | 1.03(0.84,1.26) | 0.99(0.93,1.06) | 1.03(0.96,1.10) | 1.05(0.99,1.12) | 0.92(0.83,1.01) |
| Puerto Rican vs. Dominican background | 0.17(0.02,1.24) | 1.74(0.48,6.31) | **2.66(1.01,7.01)^†^** | 1.08(0.35,3.36) | 2.24(0.91,5.47) |
| Study site – Bronx vs. Chicago | **--** | 0.85(0.30,2.45) | 1.01(0.34,3.00) | 0.52(0.19,1.43) | **4.58(1.12,18.72)^†^** |
| Born in the 50 U.S. states/D.C. | 4.92(0.59,41.06) | 2.82(0.93,8.53) | 1.31(0.49,3.52) | 1.03(0.33,3.15) | 0.79(0.26,2.41) |
| Has employer-based or private health insurance coverage | 3.23(0.74,14.19) | 0.54(0.18,1.65) | 0.43(0.18,1.00) | 1.71(0.65,4.53) | 2.44(0.88,6.78) |
| %predicted pre-bronchodilator FEV1 | 1.03(0.95,1.11) | 1.01(0.99,1.03) | 1.00(0.97,1.02) | 1.01(0.99,1.04) | 0.99(0.95,1.03) |
| Pre-bronchodilator FEV1/FVC | 0.88(0.73,1.05) | **0.88(0.83,0.94)^‡^** | **0.92(0.87,0.98)^†^** | **0.88(0.82,0.93)^‡^** | 1.05(0.98,1.14) |

COPD=chronic obstructive pulmonary disease; FEV1=forced expiratory volume in 1 second; FVC=forced vital capacity

^*^A total of 472 participants (278 controls and 194 asthma cases) were included in the analysis. Two hundred and one participants were excluded from physician-diagnosed COPD (n=45), current smokers (n=94), or former smokers with ≥10 pack-years of smoking (n=93).

^†^*P* < 0.05; ^‡^*P* < 0.01.

**eTable 2. Multinomial logistic regression analysis of asthma endotypes in SOL-Asthma (with controls as the reference outcome category), adjusting for household income instead of type of health insurance**

|  | Asthma endotypes | | | | |
| --- | --- | --- | --- | --- | --- |
| Variables | **T2^HIGH^**  **(*n*=15)** | **T1^HIGH^**  **(*n*=69)** | **T17^HIGH^**  **(*n*=93)** | **T1^HIGH^/T17^HIGH^**  **(*n*=49)** | **T2^LOW^/T1^LOW^/T17^LOW^**  **(*n*=72)** |
|  | **Odds ratio (95% confidence interval)** | | | | |
| Age (years) | 0.97(0.89,1.05) | 1.03(0.99,1.06) | 1.01(0.98,1.04) | **0.96(0.92,0.99)^†^** | 1.04(0.98,1.11) |
| Female sex | 2.29(0.58,9.05) | **2.88(1.12,7.41)^†^** | 1.90(0.85,4.23) | **3.04(1.36,6.77)^‡^** | **2.87(1.25,6.60)^†^** |
| Body mass index (kg/m^2^) | **0.83(0.69,0.99)^†^** | 1.02(0.96,1.09) | 1.06(0.99,1.13) | 1.00(0.94,1.07) | 0.97(0.89,1.04) |
| Puerto Rican vs. Dominican background | **0.17(0.04,0.75)^†^** | 0.86(0.28,2.64) | 2.29(0.95,5.56) | 1.49(0.47,4.72) | 2.07(0.74,5.77) |
| Study site – Bronx vs. Chicago | -- | 0.60(0.27,1.31) | 0.91(0.43,1.95) | 0.61(0.21,1.78) | 1.81(0.62,5.34) |
| Born in the 50 U.S. states/DC | 2.31(0.33,16.31) | **2.54(1.03,6.31)^†^** | 0.87(0.35,2.16) | 0.99(0.33,2.94) | 1.29(0.54,3.12) |
| Household income - ≥ $30,000/year vs. < $30,000/year | **6.47(1.54,27.25)^†^** | 0.44(0.16,1.21) | 0.44(0.18,1.06) | 0.64(0.25,1.63) | 0.50(0.19,1.36) |
| Smoking status |  |  |  |  |  |
| Former vs. never | 2.01(0.28,14.38) | **3.77(1.52,9.37)^‡^** | 1.96(0.90,4.29) | 0.87(0.34,2.24) | 1.58(0.58,4.35) |
| Current vs. never | 4.62(0.79,27.03) | 2.36(0.67,8.30) | 0.89(0.28,2.86) | 0.49(0.14,1.77) | **3.42(1.37,8.54)^‡^** |
| Pack-years of smoking | 0.92(0.79,1.08) | 0.98(0.96,1.01) | 0.99(0.97,1.02) | 0.98(0.95,1.01) | 0.99(0.97,1.01) |
| Physician-diagnosed COPD | 8.60(0.58,127.08) | 2.11(0.45,9.87) | **4.78(1.32,17.24)^†^** | **10.93(2.97,40.20)^‡^** | **3.25(1.10,9.67)^†^** |
| %predicted pre-bronchodilator FEV1 | 1.00(0.93,1.07) | 1.00(0.98,1.02) | 1.00(0.98,1.02) | 0.98(0.96,1.01) | 0.98(0.94,1.02) |
| Pre-bronchodilator FEV1/FVC | 0.93(0.79,1.08) | **0.93(0.88,0.98)^‡^** | **0.94(0.90,0.99)^‡^** | **0.95(0.90,0.99)^†^** | 0.98(0.91,1.06) |

COPD=chronic obstructive pulmonary disease; FEV1=forced expiratory volume in 1 second; FVC=forced vital capacity

^*^A total of 644 participants (346 controls and 302 asthma cases) were included in the analysis. Twenty-nine participants had missing data for household income.

^†^*P* < 0.05; ^‡^*P* < 0.01.

#### **eTable 3. Multinomial logistic regression analysis of asthma endotypes in SOL-Asthma (with controls as the reference outcome category), adjusting the first two genetic principal components instead of Hispanic/Latino background**

|  | Asthma endotypes | | | | | | | |
| --- | --- | --- | --- | --- | --- | --- | --- | --- |
| Variables | **T2^HIGH^**  **(*n*=14)** | **T1^HIGH^**  **(*n*=64)** | | **T17^HIGH^**  **(*n*=82)** | **T1^HIGH^/T17^HIGH^**  **(*n*=47)** | | **T2^LOW^/T1^LOW^/T17^LOW^**  **(*n*=62)** | |
|  | **Odds ratio (95% confidence interval)** | | | | | | | |
| Age (years) | **0.93(0.86,1.00)^†^** | 1.01(0.97,1.04) | 0.99(0.97,1.02) | | | **0.96(0.93,1.00)^†^** | | 1.05(1.00,1.10) |
| Female sex | 2.52(0.75,8.42) | **4.32(1.67,11.17)^‡^** | 1.99(0.86,4.60) | | | **3.74(1.47,9.54)^‡^** | | 3.75(1.54,9.13)‡ |
| Body mass index (kg/m^2^) | 1.03(0.83,1.27) | 1.01(0.95,1.08) | **1.06(1.00,1.12)^†^** | | | 1.00(0.94,1.07) | | 0.95(0.88,1.02) |
| Study site – Bronx vs. Chicago | **--** | 0.75(0.32,1.77) | 0.77(0.37,1.60) | | | 0.66(0.26,1.70) | | 2.29(0.65,8.02) |
| Born in the 50 U.S. states/DC | 2.64(0.59,11.79) | 1.83(0.84,3.97) | 0.97(0.38,2.43) | | | 0.90(0.37,2.16) | | 0.93(0.35,2.48) |
| Has employer-based or private health insurance coverage | **3.86(1.08,13.76)^†^** | 0.76(0.29,1.96) | 0.83(0.44,1.56) | | | 0.78(0.28,2.18) | | 2.22(0.94,5.23) |
| Smoking status |  |  |  | | |  | |  |
| Former vs. never | 2.04(0.21,20.07) | **4.54(1.66,12.44)^‡^** | 2.17(0.93,5.09) | | | 1.33(0.52,3.40) | | 1.49(0.55,4.05) |
| Current vs. never | 2.31(0.30,17.91) | 2.99(0.80,11.16) | 1.16(0.33,4.04) | | | 0.91(0.24,3.46) | | **5.85(2.03,16.87)^‡^** |
| Pack-years of smoking | 0.94(0.79,1.10) | 0.98(0.94,1.02) | 0.99(0.96,1.02) | | | 0.98(0.95,1.02) | | 1.00(0.98,1.02) |
| Physician-diagnosed COPD | 4.16(0.27,63.43) | 2.21(0.39,12.51) | **6.57(1.53,28.24)^†^** | | | **10.83(2.81,41.69)^‡^** | | **4.37(1.37,13.98)^†^** |
| %predicted pre-bronchodilator FEV1 | 1.05(0.97,1.15) | 0.99(0.97,1.01) | 1.01(0.99,1.03) | | | **0.97(0.95,1.00)^†^** | | 0.98(0.94,1.01) |
| Pre-bronchodilator FEV1/FVC | 0.85(0.73,1.00) | **0.93(0.88,0.98)^‡^** | **0.93(0.89,0.97)^‡^** | | | **0.95(0.91,1.00)^†^** | | 0.97(0.92,1.03) |

COPD=chronic obstructive pulmonary disease; FEV1=forced expiratory volume in 1 second; FVC=forced vital capacity

^*^A total of 580 participants (311 controls and 269 asthma cases) were included in the analysis. Ninety-three participants had missing value for genotype data were thus excluded from this analysis.

^†^*P* < 0.05; ^‡^*P* < 0.01.

#### **eTable 4. Multinomial logistic regression analysis of asthma endotypes (with T2^LOW^/T1^LOW^/T17^LOW^ asthma as the reference outcome category) and asthma severity/control at the baseline visit in SOL-Asthma**

|  | Asthma endotypes | | | |
| --- | --- | --- | --- | --- |
| Variables | **T2^HIGH^**  **(*n*=15)** | **T1^HIGH^**  **(*n*=60)** | **T17^HIGH^**  **(*n*=79)** | **T1^HIGH^/T17^HIGH^**  **(*n*=44)** |
|  | **Odds ratio (95% confidence interval)** | | | |
| Age (years) | **0.91(0.85,0.97)^‡^** | 0.96(0.91,1.02) | **0.94(0.89,1.00)^†^** | **0.91(0.85,0.97)^‡^** |
| Female sex | 0.93(0.16,5.27) | 0.78(0.17,3.53) | 0.72(0.25,2.09) | 1.02(0.26,3.96) |
| Body mass index (kg/m^2^) | 1.09(0.91,1.30) | 1.03(0.95,1.12) | 1.06(0.98,1.15) | 1.00(0.91,1.10) |
| Puerto Rican vs. Dominican background | **0.10(0.01,1.00)^†^** | 0.87(0.21,3.54) | 1.55(0.50,4.78) | 0.62(0.15,2.51) |
| Study site – Bronx vs. Chicago | **--** | 0.39(0.11,1.34) | 0.34(0.09,1.28) | **0.18(0.04,0.83)^†^** |
| Born in 50 U.S. states/DC | 2.94(0.34,25.31) | 1.23(0.42,3.65) | 0.41(0.13,1.24) | 0.39(0.09,1.61) |
| Has employer-based or private health insurance coverage | 3.19(0.89,11.47) | 0.55(0.17,1.76) | **0.36(0.13,0.98)^†^** | 0.47(0.13,1.72) |
| Smoking status – |  |  |  |  |
| Former vs. never | 0.89(0.06,14.26) | 1.95(0.53,7.12) | 0.87(0.23,3.34) | 0.27(0.06,1.18) |
| Current vs. never | 0.85(0.17,4.33) | 0.38(0.08,1.84) | **0.14(0.03,0.62)**^‡^ | **0.12(0.02,0.55)**^‡^ |
| Pack-years of smoking | 0.94(0.79,1.12) | 1.00(0.97,1.02) | 1.00(0.98,1.03) | 0.99(0.97,1.02) |
| Physician-diagnosed COPD | 3.31(0.17,63.08) | 0.52(0.10,2.72) | 1.60(0.28,9.02) | 2.49(0.52,12.03) |
| %predicted pre-bronchodilator FEV1 | 1.07(0.99,1.15) | 1.01(0.97,1.05) | 1.01(0.97,1.05) | 0.98(0.94,1.03) |
| Pre-bronchodilator FEV1/FVC | 0.89(0.77,1.04) | 0.97(0.90,1.04) | 0.98(0.91,1.05) | 1.00(0.92,1.08) |
| Asthma Control Test score | 0.85(0.72,1.01) | 0.97(0.87,1.08) | 1.04(0.93,1.16) | 1.05(0.91,1.20) |
| Current use of inhaled corticosteroids | 0.25(0.03,2.33) | 0.85(0.27,2.69) | 1.36(0.41,4.51) | 0.92(0.21,3.97) |
| Current use of oral corticosteroids | 0.75(0.11,5.22) | 0.75(0.18,3.13) | 1.06(0.34,3.27) | 0.74(0.21,2.63) |
| ≥1 ED visit or hospitalization for asthma in the prior year‡ | 3.42(0.64,18.26) | **6.95(2.44,19.80) ^‡^** | 0.95(0.28,3.19) | 1.36(0.28,6.65) |

COPD=chronic obstructive pulmonary disease; ACT=asthma control test; FEV1=forced expiratory volume in 1 second; FVC=forced vital capacity; ED=emergency department.

**^*^** A total of 259 participants with asthma were included in this analysis. We excluded participants missing adequate spirometry tests (*n*=4) or data on: ACT score, corticosteroid use, or ED visits/hospitalizations for asthma (*n*=50).

^†^*P* < 0.05; ^‡^*P* < 0.01.

#### **eTable 5. Multivariable logistic regression analysis of asthma endotypes at the baseline visit and ED visits or hospitalizations for asthma during one year of follow up in SOL-Asthma**

| Variables | Unadjusted | Adjusted^¶^ |
| --- | --- | --- |
|  | **OR (95% confidence interval)** | |
| Age (years) | 0.98(0.96,1.01) | 0.98(0.95,1.02) |
| Female sex | 1.26(0.37,4.27) | 0.92(0.29,2.94) |
| Body mass index (kg/m^2^) | 1.00(0.94,1.06) | 0.96(0.90,1.03) |
| Puerto Rican vs. Dominican background | 2.36(0.85,6.51) | 2.56(0.70,9.43) |
| Study site – Bronx vs. Chicago | 1.63(0.43,6.23) | **3.51(1.06,11.60)**^†^ |
| Born in the 50 U.S. states/DC | 1.15(0.45,2.93) | 0.57(0.20,1.66) |
| Has employer-based or private health insurance coverage | 0.77(0.25,2.40) | 1.51(0.52,4.43) |
| Smoking status – |  |  |
| Former vs. never | **3.23(1.12,9.32)**^†^ | **3.60(1.22,10.62)^†^** |
| Current vs. never | **4.85(1.55,15.24)**^†^ | **4.10(1.18,14.26)^†^** |
| Physician-diagnosed COPD | 1.88(0.48,7.34) | 1.19(0.35,4.00) |
| Inhaled corticosteroids use^§^ |  |  |
| At either visit vs. never | **3.61(1.30,10.04)**^†^ | **3.47(1.23,9.84)**^†^ |
| At both visits vs. never | **3.32(1.06,10.40)**^†^ | **3.14(1.11,8.87)**^†^ |
| %predicted pre-bronchodilator FEV1 | 0.99(0.97,1.01) | 0.97(0.94,1.00) |
| Pre-bronchodilator FEV1/FVC | 1.00(0.97,1.04) | 1.06(0.99,1.12) |
| Asthma endotypes  T2^LOW^/T1^LOW^/T17^LOW^ 1.0 1.0 | | |
| T2^HIGH^ | 0.63(0.09,4.43) | 1.24(0.09,16.52) |
| T1^HIGH^ | 2.52(0.73,8.76) | **4.31(1.26,14.7)**^†^ |
| T17^HIGH^ | 1.09(0.30,3.98) | 1.50(0.41,5.45) |
| T1^HIGH^/T17^HIGH^ | 1.64(0.30,8.94) | 3.02(0.83,11.05) |

ED=emergency department; COPD=chronic obstructive pulmonary disease; FEV1=forced expiratory volume in 1 second; FVC=forced vital capacity.

* A total of 298 participants with asthma were included in this analysis. Fifteen participants were excluded from this analysis due to missing data for inhaled corticosteroid use (n=11) or not having adequate lung function measures (n=4).

^§^ Use of inhaled corticosteroids were defined as never used in the baseline visit and 1-year follow-up visit, used in either visits, or used in both visits

¶ Model adjusted for all variables in the column.

^†^*P* < 0.05; ^‡^*P* < 0.01.

#### **eTable 6. Multivariable analysis of ACT score decline (≥3 points) during 1-year follow-up in SOL-Asthma**

| Variables | Model 1 | Model 2 |
| --- | --- | --- |
|  | **OR (95% confidence interval)** | |
| Age (years) | 0.99(0.96,1.03) | 0.99(0.95,1.03) |
| Female sex | 1.25(0.45,3.49) | 1.25(0.45,3.48) |
| Body mass index (kg/m^2^) | **0.91(0.84,0.98)**^†^ | **0.91(0.84,0.98)**^†^ |
| Puerto Rican vs. Dominican background | 1.18(0.36,3.83) | 1.12(0.37,3.38) |
| Study site – Bronx vs. Chicago | **4.23(1.62,11.01)**^‡^ | **4.47(1.63,12.29)**^‡^ |
| Born in the 50 U.S. states/DC | 0.68(0.24,1.96) | 0.68(0.23,2.00) |
| Has employer-based or private health insurance coverage | 0.91(0.34,2.47) | 0.94(0.35,2.53) |
| Smoking status – |  |  |
| Former vs. never | 2.17(0.87,5.36) | 1.94(0.76,4.95) |
| Current vs. never | 1.84(0.58,5.88) | 1.61(0.47,5.55) |
| Physician-diagnosed COPD | *NOT INCLUDED* | 2.00(0.54,7.34) |
| Inhaled corticosteroids use^§^ |  |  |
| At either visit vs. never | 2.58(0.70,9.50) | 2.69(0.75,9.63) |
| At both visits vs. never | **7.73(2.62,22.86)**^‡^ | **8.27(2.85,24.05)**^‡^ |
| ACT score at the baseline visit | **1.13(1.02,1.25)**^†^ | **1.15(1.04,1.28)**^†^ |
| Asthma endotypes | | |
| T2^LOW^/T1^LOW^/T17^LOW^ | 1.0 | 1.0 |
| T2^HIGH^ | 0.49(0.07,3.22) | 0.48(0.08,3.09) |
| T1^HIGH^ | 1.27(0.39,4.13) | 1.33(0.40,4.42) |
| T17^HIGH^ | 1.44(0.55,3.76) | 1.44(0.54,3.85) |
| T1^HIGH^/T17^HIGH^ | 1.93(0.66,5.62) | 1.77(0.62,5.06) |

COPD=chronic obstructive pulmonary disease; ACT=asthma control test

*A total of 301 participants with asthma were included in this analysis (as 12 participants were excluded due to missing data on inhaled corticosteroid use or ACT score at either visit).

^§^ Use of inhaled corticosteroids was defined as never used, used at the baseline visit or the 1-year follow-up visit, or used at both visits

^†^*P* < 0.05; ^‡^*P* < 0.01.

#### **eTable 7. Multivariable analysis of change in lung function between the baseline visit in the Hispanic Community Health Study/Study of Latinos (HCHS/SOL) and the baseline visit in SOL-Asthma**

|  | Change in lung function measures^*^  β (95% confidence interval) | | |
| --- | --- | --- | --- |
| Variables | **Δ %predicted pre-bronchodilator FEV1** | **Δ %predicted pre-bronchodilator FVC** | **Δ pre-bronchodilator FEV1/FVC** |
| Body mass index (kg/m^2^) | **-0.29(-0.52,-0.06)**^†^ | **-0.48(-0.67,-0.29)**^‡^ | **0.12(0.02,0.23)**^†^ |
| Puerto Rican vs. Dominican background | -1.18(-3.84,1.47) | -0.42(-2.97,2.12) | -0.49(-2.07,1.09) |
| Study site – Bronx vs. Chicago | 1.93(-1.26,5.11) | 2.65(-0.33,5.62) | -0.40(-1.74,0.93) |
| Born in the 50 U.S. states or D.C. | 0.97(-2.24,4.18) | 1.38(-1.13,3.89) | -0.35(-2.13,1.43) |
| Has employer-based or private health insurance coverage | 1.41(-0.99,3.81) | 1.04(-1.40,3.48) | 0.32(-0.68,1.31) |
| Smoking status |  |  |  |
| Former vs. never | 1.24(-1.58,4.07) | 1.62(-0.65,3.88) | -0.51(-1.88,0.87) |
| Current vs. never | 0.80(-2.26,3.86) | 1.51(-1.25,4.28) | -0.43(-2.17,1.30) |
| Pack-years of smoking | -0.03(-0.09,0.03) | -0.02(-0.07,0.02) | -0.001(-0.03,0.03) |
| Physician-diagnosed COPD | -5.17(-10.73,0.38) | -1.77(-7.20,3.66) | **-4.15(-6.97,-1.33)**^‡^ |
| Corresponding baseline measure | **-0.26(-0.34,-0.17)**^‡^ | **-0.29(-0.36,-0.21)**^‡^ | **-0.19(-0.29,-0.10)**^‡^ |
| Time interval between two visits | 0.87(-0.34,2.09) | 0.75(-0.39,1.89) | -0.07(-0.58,0.44) |
| Asthma endotypes (vs. controls) |  |  |  |
| T2^HIGH^ | -1.30(-7.93,5.33) | 1.44(-4.69,7.57) | -2.03(-4.45,0.39) |
| T1^HIGH^ | -0.96(-5.13,3.21) | 2.31(-0.99,5.61) | **-1.99(-3.82,-0.16)**^†^ |
| T17^HIGH^ | -1.08(-5.16,3.01) | 0.88(-2.32,4.09) | -1.46(-3.48,0.55) |
| T1^HIGH^ /T17^HIGH^ | -2.38(-5.87,1.10) | -0.61(-3.99,2.78) | **-2.06(-3.76,-0.35)**^†^ |
| T2^LOW^/T1^LOW^/T17^LOW^ | 0.11(-3.15,3.37) | 2.87(-0.28,6.02) | **-2.29(-4.03,-0.54)**^†^ |

COPD=chronic obstructive pulmonary disease; FEV1=forced expiratory volume in 1 second; FVC=forced vital capacity

^*^Participants with lung function measures that were missing (n=40) or had inadequate quality (n=36) at either visit (*n*=40) were excluded, leaving 597 SOL-Asthma participants in this analysis.

^†^*P* < 0.05; ^‡^*P* < 0.01.

#### **eTable 8. Main characteristics of 353 participants in the CAAPA, by asthma status**

|  | Without asthma  (controls, *n*=191) | With asthma (cases, *n*=162) | | | | |
| --- | --- | --- | --- | --- | --- | --- |
| Characteristics |  | **T2^HIGH^**  **(*n*=25)** | **T1^HIGH^**  **(*n*=45)** | **T17^HIGH^**  **(*n*=21)** | **T1^HIGH^/T17^HIGH^**  **(*n*=27)** | **T2^LOW^/T1^LOW^/T17^LOW^**  **(*n*=44)** |
| Age, years | 38.8±13.4 | 37.4±13.4 | 38.0±13.2 | 35.9±10.1 | 40.8±12.2 | **45.7±16.1**^*^ |
| Female sex | 121(63.4) | 16(64.0) | 33(73.3) | 18(66.7) | 16(76.2) | 28(63.6) |
| Body mass index (kg/m^2^) | 28.3±6.0 | 27.8±5.6 | **31.2±7.4**^*^ | 29.6±6.8 | **32.0±6.0**^*^ | 30.2±7.4 |
| Study site |  |  |  |  |  |  |
| In the U.S. | 97(50.8) | 3(12.0) | 21(46.7) | 2(9.5) | 18(66.7) | 24(54.5) |
| Outside the U.S. | 94(49.2) | **22(88.0)**^*^ | 24(53.3) | **19(90.5)**^*^ | **9(33.3)**^*^ | 20(45.5) |
| Smoking status |  |  |  |  |  |  |
| Never | 175(92.1) | 24(96.0) | 38(84.4) | 20(100.0) | 25(92.6) | 38(90.5) |
| Former | 15(7.9) | 1(4.0) | 7(15.6) | 0(0.0) | 2(7.4) | 4(9.5) |
| CASI score | **--** | 4.6±3.6 | 4.7±3.4 | 6.7±4.2 | 4.4±3.8 | 4.5±3.2 |
| Current use of inhaled corticosteroids | **--** | 11(44.0) | 22(48.9) | 13(61.9) | 12(44.4) | 17(38.6) |
| Current use of oral corticosteroids or hospitalizations for asthma | **--** | 4(16.0) | 7(15.6) | 6(28.6) | 3(11.1) | 8(18.2) |
| %predicted pre-bronchodilator FEV1 | 93.0±12.9 | **73.4±23.0**^*^ | **81.5±24.1**^*^ | **63.1±8.6**^*^ | **79.2±24.9**^*^ | **77.7±18.4**^*^ |
| %predicted pre-bronchodilator FVC | 93.9±13.8 | **80.2±22.4**^*^ | 90.9±18.1 | **75.1±8.8**^*^ | 91.3±19.4 | 88.8±16.6 |
| Pre-bronchodilator FEV1/FVC (%) | 82.2±5.7 | **74.8±10.6**^*^ | **72.8±10.8**^*^ | **69.9±8.7**^*^ | **75.6±8.8**^*^ | **71.0±10.6**^*^ |

CAAPA: Consortium on Asthma among African ancestry Populations in the Americas; CASI: Composite Asthma Severity Index; FEV1: forced expiratory volume in 1 second; FVC: forced vital capacity.

Data are shown as n (%) for categorical variables and as mean (standard deviation) for continuous variables. Numbers may vary due to missingness.

^*^*P*<0.05 for comparison of participants within each asthma endotype versus controls or (for outcomes occurring only in cases) participants across asthma endotypes.

#### **eTable 9. Multinomial regression analysis of asthma endotypes (with controls as the reference outcome category) in CAAPA**

|  | Asthma endotypes | | | | |
| --- | --- | --- | --- | --- | --- |
| Variables | **T2^HIGH^**  **(*n*=25)** | **T1^HIGH^**  **(*n*=45)** | **T17^HIGH^**  **(*n*=21)** | **T1^HIGH^/T17^HIGH^**  **(*n*=27)** | **T2^LOW^/T1^LOW^/T17^LOW^**  **(*n*=44)** |
|  | **Odds ratio (95% confidence interval)** | | | | |
| Age (years) | 0.98 (0.93, 1.03) | 0.96 (0.93, 1.00) | **0.92 (0.86, 0.99)**^†^ | 1.00 (0.96, 1.05) | 1.03 (1.00, 1.07) |
| Female sex | 1.36 (0.4, 4.57) | 2.01 (0.73, 5.5) | 2.12 (0.53, 8.44) | 0.97 (0.35, 2.71) | 1.09 (0.41, 2.90) |
| Body mass index (kg/m^2^) | 1.00 (0.90, 1.10) | **1.07 (1.00, 1.14)**^†^ | 1.02 (0.92, 1.13) | 1.08 (1.00, 1.16) | 1.03 (0.96, 1.10) |
| Study site – Outside the U.S. vs. In the U.S. | **5.40 (1.10, 26.36)**^†^ | 1.45 (0.44, 4.74) | **5.94 (1.02, 34.74)**^†^ | **0.18 (0.05, 0.68)**^†^ | **0.22 (0.06, 0.76)** ^†^ |
| Smoking status – Never vs. Former | 0.85 (0.08, 9.45) | 0.41 (0.10, 1.64) | **--** | 2.43 (0.41, 14.31) | 2.28 (0.51, 10.18) |
| %predicted pre-bronchodilator FEV1 | 0.96 (0.92, 1.01) | 1.01 (0.98, 1.05) | **0.92 (0.87, 0.97)**^‡^ | **0.95 (0.92, 0.99)**^‡^ | 0.97 (0.94, 1.00) |
| Pre-bronchodilator FEV1/FVC (%) | **0.90 (0.83, 0.98)**^†^ | **0.81 (0.76, 0.88)**^‡^ | **0.91 (0.84, 0.99)**^†^ | **0.91 (0.84, 0.97)**^‡^ | **0.84 (0.79, 0.91)**^‡^ |

CAAPA: Consortium on Asthma among African ancestry Populations in the Americas; CASI: Composite Asthma Severity Index; FEV1: forced expiratory volume in 1 second; FVC: forced vital capacity.

^*^ Additionally adjusted for the first two genetic principal components derived from genome-wide genotypic data.

^†^*P* < 0.05; ^‡^*P* < 0.01.
